# Removing animal and nonhuman records in Ovid Embase: A comparison of 11 filters

**DOI:** 10.64898/2026.02.13.26346239

**Authors:** Helen Fulbright, Connor Evans

**Affiliations:** MCLIP Information Specialist / Research Fellow in Information Science, Centre for Reviews and Dissemination, University of York, York, United Kingdom; Research Fellow in Evidence Synthesis, Centre for Reviews and Dissemination, University of York, York, United Kingdom

**Keywords:** bibliographic database, Embase, MEDLINE, indexing, information retrieval, search strategies

## Abstract

**Introduction:** Several filters are routinely used to remove animal or nonhuman records in Ovid Embase, despite there being no performance data for them. The filters take different approaches in design.

**Objective:** To understand and compare the impact of 11 filters to remove animal or nonhuman records in Ovid Embase. To understand the indexing of relevant subject headings in Embase.

**Methods:** To assess filter performance, we screened and categorised 3,000 records as ‘should be removed’ or ‘should be retained’ and calculated the sensitivity, specificity and overall accuracy for each filter. We reported on the focus or content of records that were incorrectly removed, using seven categories.

**Results:** Method 11 was the most sensitive, correctly retaining 90.6% records, whereas method 3 had the highest specificity, correctly removing 71.5% records. Out of seven categories, those in category 1 ‘uses human participants or data’ were the most excluded.

**Discussion:** Filters that did not remove nonhuman records had higher sensitivity. Filter performance could vary by subject, publication type and language due to differences in indexing.

**Conclusion:** In choosing a search filter, information specialists and review teams should discuss whether animals or nonhumans could feature in relevant studies.

## INTRODUCTION

Evidence syntheses interested in humans as a subject or population often remove records on animals and nonhumans where certain studies (e.g., pre-clinical studies using animals) are ineligible for inclusion. These records are removed using search filters - pre-tested search terms designed to filter results to specific topics, populations, research methods, or study types [1]. Filters described as ‘validated’ have been tested against a ‘gold standard’ or set of known relevant and irrelevant records, whereas ‘non-validated’ filters may have undergone limited testing [2]. Several search filters without performance data are routinely used to remove animal or nonhuman records in Ovid Embase. Although the database contains an inbuilt limit to filter to results indexed with the Emtree subject heading ‘human/’, this risks exclusion of relevant records without it. The various filters designed for Ovid Embase mainly follow the same structure as a method designed for Ovid MEDLINE, which is applied in the Cochrane highly-sensitive search strategies for randomised controlled trials (RCTs) [4]. The filter removes records indexed with ‘exp animals/’ that are not also indexed with ‘humans/’. It is designed to limit records to those likely to be about humans by excluding records that are *only* about animals [3].

A much wider variety of filters to remove animal or nonhuman records are used in Ovid Embase compared with Ovid MEDLINE. Various filters of unknown authorship can be found in the appendices of published literature reviews, such as one used by the Cochrane Common Mental Disorders Group [5] and another used by York Health Economics Consortium [6]. Some are found in the strategies used to populate the Database of Abstracts of Reviews of Effects (DARE) and the NHS Economic Evaluations Database (NHS EED) [7]. Others feature in filter resources by the InterTASC Information Specialists’ Sub-Group [8] and Canada’s Drug Agency [9]. Ovid Embase contains its own inbuilt limits to remove animal or nonhuman records [10]. One is part of a validated filter to find RCTs in Ovid

Embase [11], though we do not have separate data on its performance. Filters to remove animal or nonhuman records, including the frequently used MEDLINE filter, are not routinely credited in published evidence syntheses. This reflects that the filters may be custom-made, unpublished, or non-validated and means we cannot fully understand their exact intended purpose or the context in which they were developed. Soudant and colleagues have questioned the reliability of non-validated filters to remove animal records [12].

Finnegan et al have highlighted various challenges defining and removing animal records [13]. These challenges, such as how we define an animal or nonhuman record, mean that understanding the efficacy of filters designed to remove them yields numerous complexities. Firstly, just as we cannot rely on all records that are relevant to humans to be indexed with ‘human/’, we also cannot rely on records relevant to both animals/nonhumans *and* humans to have this indexing term. Therefore, these records could be excluded from search results if desired Emtree terms such as ‘human/’ or ‘human experiment/’, have not been applied to records (i.e., are missing on records with humans as a subject or population). This is important to consider, as animals are involved in human health in a variety of ways: in animal-assisted therapies; in detecting conditions such as cancer or low blood sugar; as sources of disease transmission or injury; and in numerous other respects. Similarly, as the Emtree term ‘nonhuman/’ can encompass animals, bacteria, viruses, and plants, we may not consider the full breadth of this term during filter selection, despite the potential relevance of nonhumans to human health. This prompts us to consider several questions: how are indexers assigning Emtree headings to records; what are information specialists using the filters to remove; what types of animal or nonhuman records do research teams want removed (and how might this vary on a review-by-review basis); and do information specialists and review teams discuss whether the use of a filter is appropriate?

There are numerous differences in the animal/nonhuman filters designed for Ovid Embase. Many exclusively use Emtree terms but some also use the title and/or abstract fields. There are also several differences in approach between the filters. As information specialists using Embase have a choice of filters to remove animal or nonhuman records, it would be valuable to understand how these perform, compare to each other, and the focus or content of records they exclude incorrectly, to inform discussion and decision-making about filter use.

## OBJECTIVES

This is an exploratory study which aims to investigate the impact of 11 filters to remove animal or nonhuman records in Ovid Embase. The study intends to explore how the filters work and compare to each other to help information specialists make an informed decision about filter use. The main objectives are to:

- understand how records are indexed with Emtree subject headings and how this could affect filter performance;
- explore how the 11 filters compare in terms of the proportion of records correctly removed or retained; and
- explore which types of record were incorrectly removed according to the focus or content of the record.

## METHODS

A summary of the methodology is as follows, we:

1. communicated with Elsevier to ask questions on how records are indexed with Emtree subject headings;
2. double screened a sample of 3,000 records to judge whether records should be removed or retained, using three separate datasets; and
3. determined how the filters compare in their recall of records in the ‘should be removed’ and ‘should be retained’ categories; and in the focus or content of the records they incorrectly removed.

### 1. Understanding how records are indexed with Emtree terms

Elsevier’s 2026 indexing guide for Embase was used to understand how indexing might affect filter performance. We also e-mailed Elsevier to enquire: if indexers at Elsevier use the scope notes in determining whether to use Emtree subject headings relating to humans, animals, and nonhumans; and the meaning and interpretation of the scope notes for the Emtree term ‘human/’ [14].

Embase records are automatically indexed with Emtree terms based on their titles, abstracts and author keywords [14]. However, records which meet certain criteria (according to an automated process), are then manually indexed using the full text by indexers at Elsevier with a background in biomedicine [14]. This occurs for records with one or more Emtree terms for diseases, drugs, or devices; and records identified as: reviews; short surveys (for case reports only); meta-analyses; and systematic reviews [14]. During manual indexing, indexers at Elsevier use Emtree’s taxonomy as well as some additional rules and internal notes to ensure consistency [15].

Automatic indexing is retained for pre-prints and conference abstracts, and records with the status ‘in press’ or ‘in process’ unless listed as ‘published’ or ‘processed’ respectively [14]. This means there are differences in the indexing of conference abstracts and pre-prints in comparison to other publication types commonly sought in evidence syntheses. Records published in languages other than English are indexed based on the English title and abstract only [14] and do not meet the criteria for manual indexing, except for those in the Embase French Local Literature Module [16]. The scope notes for the Emtree heading ‘human/’, includes wording to describe its application to records ‘where humans are a feature’. Elsevier explained that this could be ‘anything from a human patient in a case report to a rural area, from a major clinical study (human subjects) to an environmental study in an urban area’ [17]. The wording was chosen ‘to include situations where there is a human presence, without the study necessarily including human subjects’ [17].

### 2. Screening records on: animals/nonhumans; humans; and animals/nonhumans and humans

To create our datasets, we developed three search strategies for Ovid Embase designed to find records likely to belong to one of the following categories:

1. animals/nonhumans;
2. humans; and
3. animals/nonhumans and humans.

These categories were chosen to provide a more comprehensive set of records than might be retained or removed by the filters compared with searching for topics only relevant to humans. Results for datasets 1 and 3 are not individually reported in this paper but are available in the supplementary material. This study focuses on data from the humans dataset (as most representative of topics that would use a filter) as well as all three datasets combined.

Each of the three search strategies included 16 lines of search terms and applied the ‘all fields’ field code (af). We chose search terms to find records likely to belong to each category only (e.g., veterinary parasitology for the animals/nonhumans category; personality disorder for the humans category; and human-animal bond for the animals/nonhumans and humans category).

The search strategies employed a filter for systematic reviews [7] as well as a filter for RCTs [11], as these study types are commonly sought in evidence syntheses. The study filters were adapted to exclude any lines that removed animal or nonhuman records. The 11 filters (see Table 1) were then employed consecutively in each of the three search strategies. These were applied with the NOT operator and the AND operator, so we could export records that would be removed or retained by the 11 methods. All lines removing or retaining records were pooled together using the OR operator. For each of the searches, we exported a sample of 500 records removed by the filters and 500 retained by the filters, totalling 1,000 for each category and 3,000 overall. Many of the 11 filters were either known to or used within the Centre for Reviews and Dissemination except for methods 8 and 9, which were adapted from method 7 for testing purposes. All search strategies are available in the appendix.

**Table 1:**
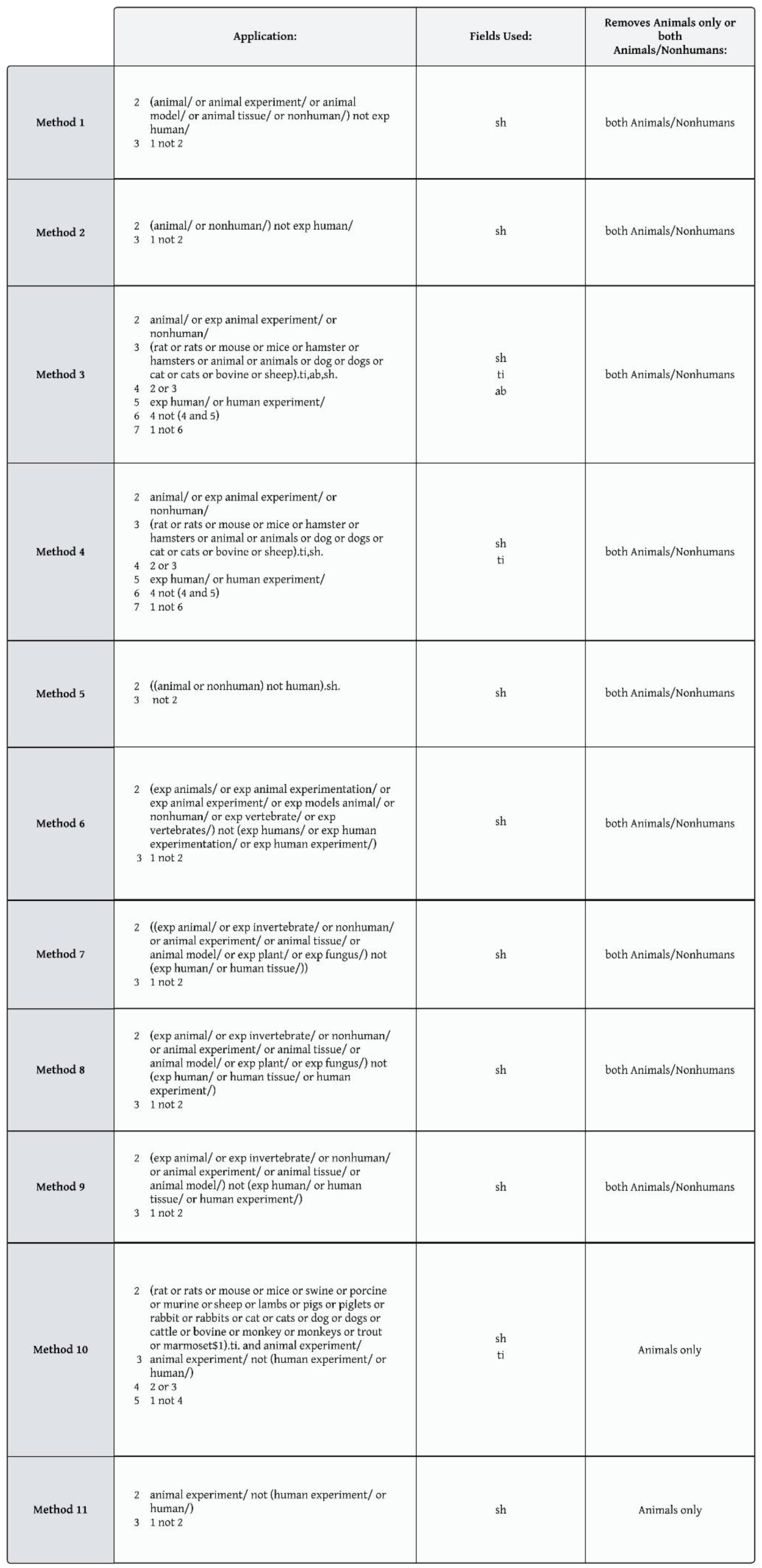
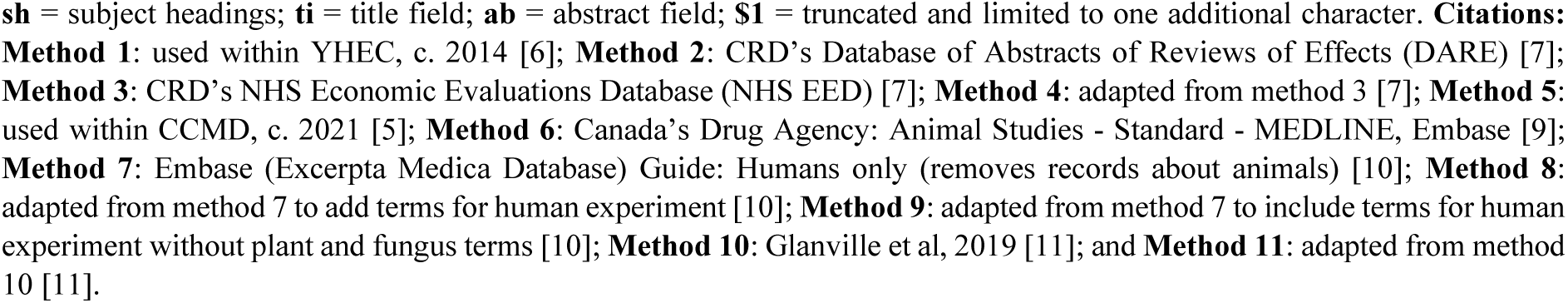
Methods of removing animal or nonhuman records in Ovid Embase.

Each record was screened by both reviewers using the titles and abstracts. Records were categorised as either ‘should be removed’ or ‘should be retained’, using the criteria and definitions in Table 2, with disagreements resolved through discussion. The definitions and screening criteria in Table 2 are subjective and were designed around the context of the record rather than the population, as this is the broadest and most sensitive approach. We chose this approach as definitions of animal/nonhuman records that are ineligible and should be excluded (both in the searches and during screening) could vary depending on the review being conducted.

**Table 2:** Definitions and screening criteria.

|  | Definitions: | Screening Criteria: |
| --- | --- | --- |
| **Animal or Nonhuman Records** | • The primary focus or outcome is *on, about, or for* animals or nonhumans, **and** there is no relevance, impact or implication for human health (i.e., studies where humans are mentioned or compared in the study would be excluded from this categorisation if there is no relevance, impact or implication for human health).<br>• Pre-clinical studies investigating human health conditions by experimenting on animals whether wholly or in part (e.g., tissues, cells, etc), including studies also using human tissues or cells (unless human tissues, cells etc are tested separately and compared with animal tissues, cells etc).<br>• Studies that feature or use animals or nonhumans whether wholly or in part (e.g., tissues, cells, etc) with no relevance, impact or implication for human health, even if humans are mentioned or whole, living humans are compared (e.g., interns versus specialists).<br>• Studies on the properties, cultivation, use, health or diseases of nonhuman organisms such as plants, fungi, viruses, bacteria etc, where there is no relevance, impact or implication for human health. | **Should be Removed:**<br>Meets criteria for animal/nonhuman records. |
| **Not Animal or Nonhuman Records** | Anything that does not meet the definition of an animal or nonhuman record above. Several examples include (but are not limited to):<br>• The focus is not *on or about* humans, animals or nonhumans (e.g., computers, the environment, etc).<br>• First-in-human studies.<br>• The primary focus or outcome is *on, about, or for* human health (except for pre-clinical studies investigating human health conditions by experimenting on animals). *On, about, or involving* animals/nonhumans or both animals/nonhumans **and** humans, where the focus or outcome is either split between humans and animals/nonhumans, or primarily about humans (e.g., training dogs to detect cancer in humans; studies of zoonotic diseases contagious to humans; nutritional value of animal products for humans). | **Should be Retained:**<br>Meets criteria for Not animal/nonhuman records. |

Table 1 shows 11 different methods for removing animal or nonhuman records in Ovid Embase. Line 1 (not listed) is representative of the line to which the method should be applied.

### 3. Testing and comparing the performance of 11 filters

To test and compare the performance of the 11 filters, we needed to find each filter’s recall of records that we allocated to the ‘should be removed’ or ‘should be retained’ categories during screening. This was performed for each dataset individually. Using the ‘an’ field code, we searched for the accession numbers of each record allocated to the ‘should be removed’ and ‘should be retained’ categories with the 11 filters applied consecutively. This was performed separately for each category. The filters were applied with the NOT operator as well as with the AND operator, so we could determine the exact number of records removed or retained by each filter. Due to duplicate removal and changes to the records since their original export from Embase, some of the 3,000 records could not be retrieved using the accession number, resulting in only 2,689 records being tested overall.

For each of the 11 filters, we calculated the sensitivity, specificity, and the overall accuracy. Sensitivity, which is a measure of a filter’s ability to correctly include relevant records, was calculated by dividing the number of records retained by each filter by the number of records in the ‘should be retained’ category and then multiplying this number by 100. Specificity, which is a measure of a filter’s ability to correctly exclude irrelevant records, was calculated by dividing the number of records removed by each filter by the number of records in the ‘should be removed’ category and then multiplying this number by 100. In this paper, the term ‘overall accuracy’ is used to refer to the number of records that were either correctly removed or correctly retained by each filter. This was calculated by combining the number of correctly retained and removed records for each method, dividing this number by the combined number of records that should be removed or retained, and then multiplying this number by 100. Overall accuracy should be understood using the proportion of correct records in the ‘should be retained’ and ‘should be removed’ categories, as this weights the overall accuracy in favour of either sensitivity or specificity accordingly. For all calculations, percentages are shown to one decimal point.

All 1,091 records in the ‘should be retained’ category were screened on title and abstract by one reviewer and assigned to one of seven categories concerning the focus or content of the record. Records were screened using the order listed below, with the earliest categorisation given to records. The categories applied were:

1. uses human participants or data
2. review or overview of a condition or intervention affecting humans
3. review or overview of a factor affecting human health
4. study outcomes relevant to diagnosing or treating humans
5. study outcomes relevant to a factor affecting human health
6. not on animals/nonhumans or human health directly
7. other (this category includes publication types such as corrections and notes)

We then created search strings of the accession numbers of records in each of the seven categories to find the number of records incorrectly removed by each filter. The 11 filters were applied consecutively in each of the search strategies. Only 1,088 of the 1,091 records could be retrieved using the accession number.

The process of testing the performance of the 11 filters is summarised in Figure 1.

**Figure 1:**
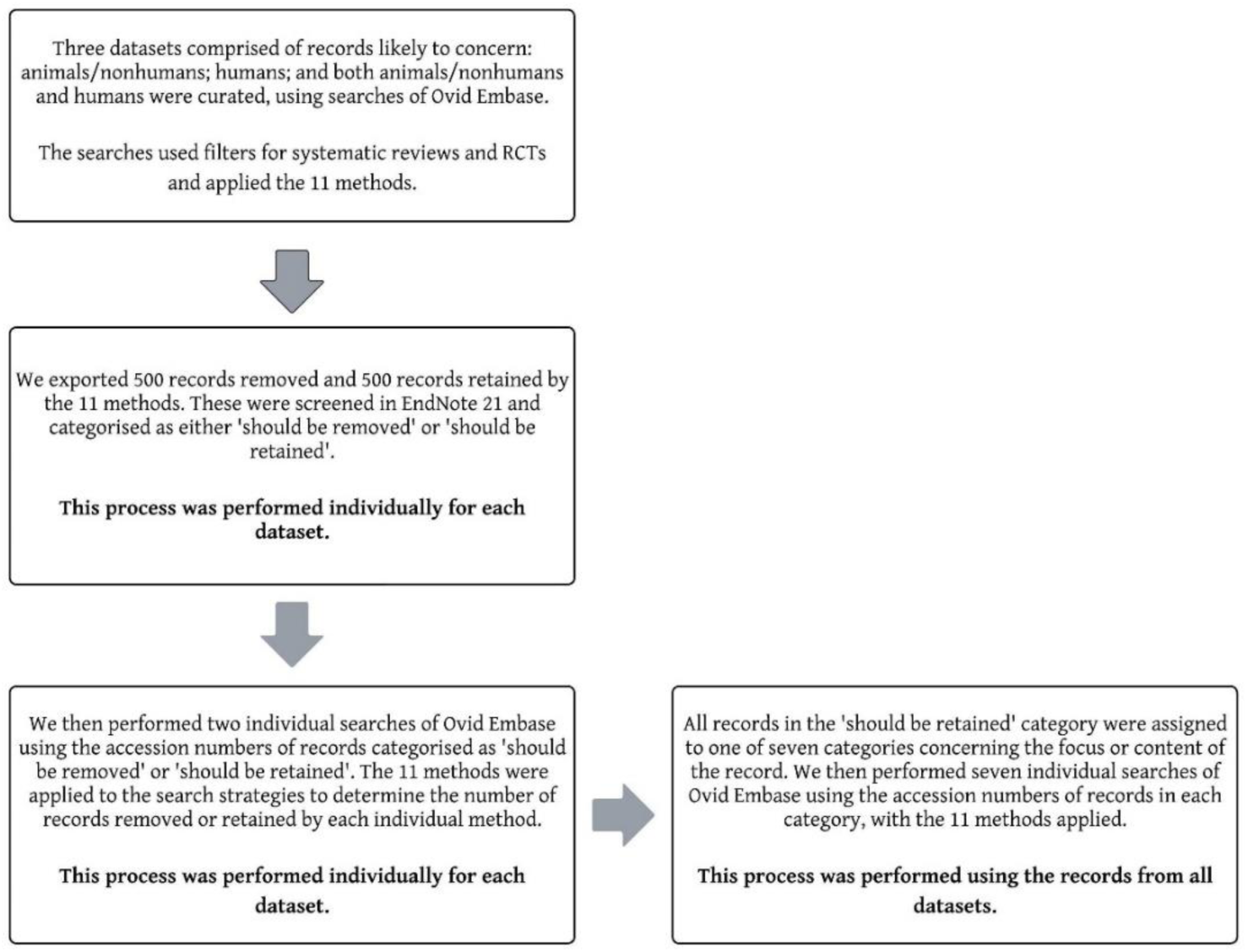
Testing the performance of 11 filters to remove animal or nonhuman records.

## RESULTS

Table 3 shows the recall of records in the ‘should be removed’ or should be retained’ categories by each of the 11 filters for the humans dataset. Results with the highest percentages in each column are shown in grey and bold.

**Table 3:** Comparing the performance of 11 filters to remove animal or nonhuman records (Humans Dataset)

| **Method:** | *Sensitivity*<br>**Should be Retained:** | *Specificity*<br>**Should be Removed:** | **Overall Accuracy:** |
| --- | --- | --- | --- |
| **1** | 531 of 678<br>(78.3%) | 281 of 312<br>(90.1%) | 812 of 990<br>(82%) |
| **2** | 532 of 678<br>(78.5%) | 281 of 312<br>(90.1%) | 813 of 990<br>(82.1%) |
| **3** | 511 of 678<br>(75.4%) | **286 of 312**<br>**(91.7%)** | 797 of 990<br>(80.5%) |
| **4** | 530 of 678<br>(78.2%) | 282 of 312<br>(90.4%) | 812 of 990<br>(82%) |
| **5** | 531 of 678<br>(78.3%) | 281 of 312<br>(90.1%) | 812 of 990<br>(82%) |
| **6** | 529 of 678<br>(78%) | 283 of 312<br>(90.7%) | 812 of 990<br>(82%) |
| **7** | 520 of 678<br>(76.7%) | 285 of 312<br>(91.3%) | 805 of 990<br>(81.3%) |
| **8** | 520 of 678<br>(76.7%) | 285 of 312<br>(91.3%) | 805 of 990<br>(81.3%) |
| **9** | 530 of 678<br>(78.2%) | 281 of 312<br>(90.1%) | 811 of 990<br>(81.9%) |
| **10** | 589 of 678<br>(86.9%) | 251 of 312<br>(80.4%) | **840 of 990**<br>**(84.8%)** |
| **11** | **599 of 678**<br>**(88.3%)** | 229 of 312<br>(73.4%) | 828 of 990<br>(83.6%) |

Method 11 was the most sensitive filter for this dataset, correctly retaining 88.3% records in the ‘should be retained’ category. A similar proportion was correctly retained by method 10 (86.9%), whereas all other filters retained similar amounts to each other, ranging between 75.4% and 78.5%.

Method 3 had highest specificity, correctly removing 91.7% records in the ‘should be removed’ category. All other filters correctly removed similar proportions of records (ranging from 90.1% to 91.3%), except for method 11 which had the lowest specificity at 73.4% and method 10 with second-lowest specificity at 80.4%.

Method 10 achieved 84.8% overall accuracy for the humans dataset. Similar percentages were observed for the other filters, which ranged from 80.5% to 83.6%.

Table 4 shows the recall of records in the ‘should be retained’ and ‘should be removed’ categories by each of the 11 filters for all datasets combined. Results with the highest percentages in each column are shown in grey and bold.

**Table 4:** Comparing the performance of 11 filters to remove animal or nonhuman records (All Datasets)

| All Datasets | Method: | *Sensitivity*<br>Should be Retained: | *Specificity*<br>Should be Removed: | Overall Accuracy: |
| --- | --- | --- | --- | --- |
|  | 1 | 868 of 1102 (78.8%) | 1079 of 1587 (68%) | 1947 of 2689 (72.4%) |
|  | 2 | 870 of 1102 (78.9%) | 1079 of 1587 (68%) | 1949 of 2689 (72.5%) |
|  | 3 | 829 of 1102 (75.2%) | **1134 of 1587 (71.5%)** | **1963 of 2689 (73%)** |
|  | 4 | 867 of 1102 (78.7%) | 1085 of 1587 (68.4%) | 1952 of 2689 (72.6%) |
|  | 5 | 869 of 1102 (78.9%) | 1080 of 1587 (68.1%) | 1949 of 2689 (72.5%) |
|  | 6 | 863 of 1102 (78.3%) | 1086 of 1587 (68.4%) | 1949 of 2689 (72.5%) |
|  | 7 | 855 of 1102 (77.6%) | 1087 of 1587 (68.5%) | 1942 of 2689 (72.2%) |
|  | 8 | 855 of 1102 (77.6%) | 1087 of 1587 (68.5%) | 1942 of 2689 (72.2%) |
|  | 9 | 865 of 1102 (78.5%) | 1083 of 1587 (68.2%) | 1948 of 2689 (72.4%) |
|  | 10 | 973 of 1102 (88.3%) | 792 of 1587 (49.9%) | 1765 of 2689 (65.6%) |
|  | 11 | **998 of 1102 (90.6%)** | 654 of 1587 (41.2%) | 1652 of 2689 (61.4%) |

Method 11 was the most sensitive filter for all datasets, correctly retaining 90.6% records in the ‘should be retained’ category. A similar proportion was correctly retained by method 10 (88.3%), whereas all other filters retained similar amounts to each other, ranging from 75.2% to 78.9%.

Method 3 had highest specificity, correctly removing 71.5% records in the ‘should be removed’ category. All other filters correctly removed similar proportions of records (ranging from 68% to 68.5%), except for method 11 which had the lowest specificity at 41.2% and method 10 with second-lowest specificity at 49.9%.

Method 3 achieved the highest overall accuracy at 73%. Similar percentages were observed for all other filters, (ranging from 72.2% to 72.6%), except for method 11 with 61.4% overall accuracy and method 10 with 65.6%.

For all datasets combined, Table 5 shows the number of incorrectly removed records by each filter and for each category. The mean percentages are listed for each filter.

**Table 5:**
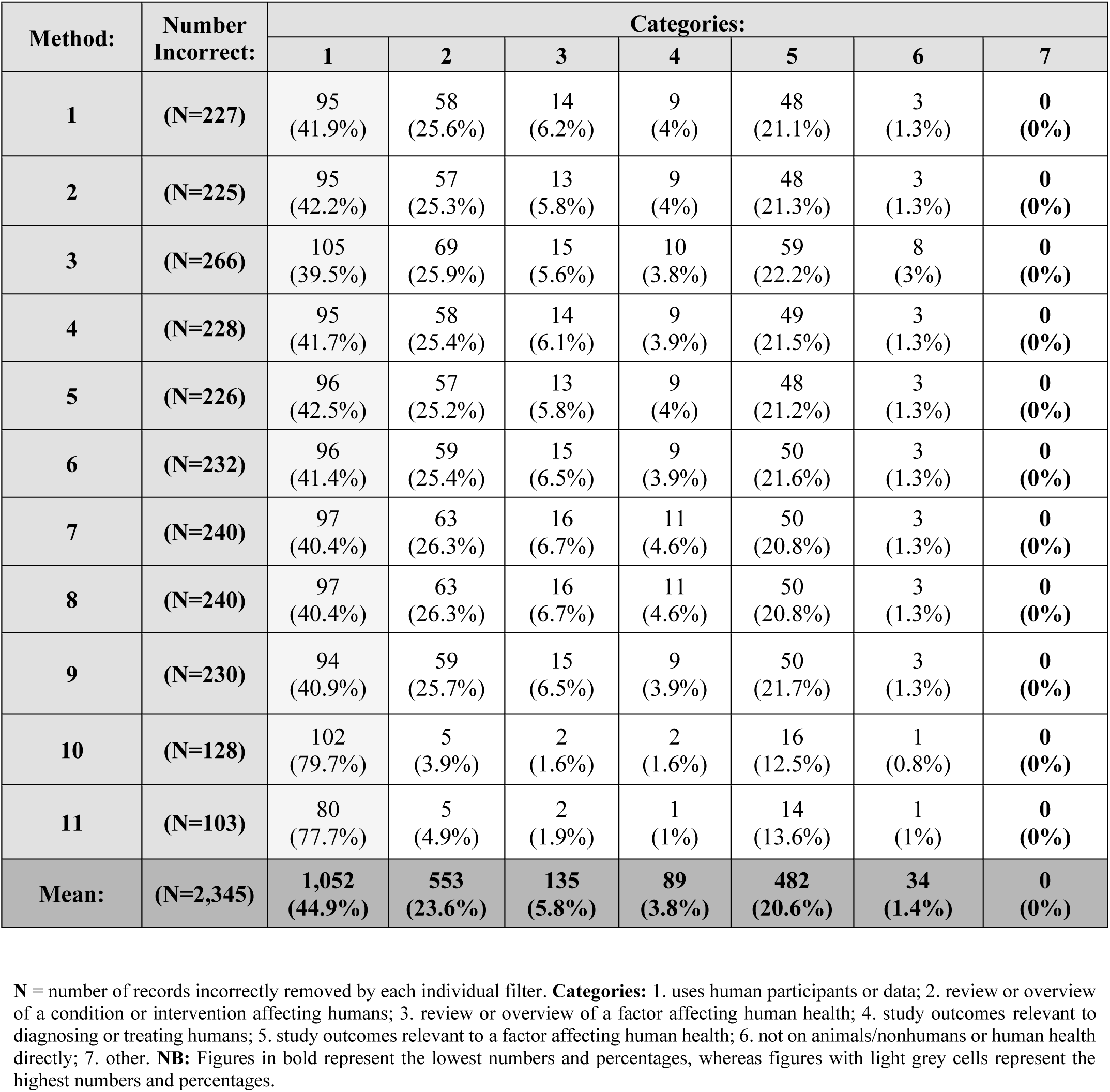
Incorrectly removed records by filter and category (All Datasets)

| Method: | Number Incorrect: | Categories: |  |  |  |  |  |  |
| --- | --- | --- | --- | --- | --- | --- | --- | --- |
|  |  | 1 | 2 | 3 | 4 | 5 | 6 | 7 |
| **1** | **(N=227)** | 95<br>(41.9%) | 58<br>(25.6%) | 14<br>(6.2%) | 9<br>(4%) | 48<br>(21.1%) | 3<br>(1.3%) | **0**<br>**(0%)** |
| **2** | **(N=225)** | 95<br>(42.2%) | 57<br>(25.3%) | 13<br>(5.8%) | 9<br>(4%) | 48<br>(21.3%) | 3<br>(1.3%) | **0**<br>**(0%)** |
| **3** | **(N=266)** | 105<br>(39.5%) | 69<br>(25.9%) | 15<br>(5.6%) | 10<br>(3.8%) | 59<br>(22.2%) | 8<br>(3%) | **0**<br>**(0%)** |
| **4** | **(N=228)** | 95<br>(41.7%) | 58<br>(25.4%) | 14<br>(6.1%) | 9<br>(3.9%) | 49<br>(21.5%) | 3<br>(1.3%) | **0**<br>**(0%)** |
| **5** | **(N=226)** | 96<br>(42.5%) | 57<br>(25.2%) | 13<br>(5.8%) | 9<br>(4%) | 48<br>(21.2%) | 3<br>(1.3%) | **0**<br>**(0%)** |
| **6** | **(N=232)** | 96<br>(41.4%) | 59<br>(25.4%) | 15<br>(6.5%) | 9<br>(3.9%) | 50<br>(21.6%) | 3<br>(1.3%) | **0**<br>**(0%)** |
| **7** | **(N=240)** | 97<br>(40.4%) | 63<br>(26.3%) | 16<br>(6.7%) | 11<br>(4.6%) | 50<br>(20.8%) | 3<br>(1.3%) | **0**<br>**(0%)** |
| **8** | **(N=240)** | 97<br>(40.4%) | 63<br>(26.3%) | 16<br>(6.7%) | 11<br>(4.6%) | 50<br>(20.8%) | 3<br>(1.3%) | **0**<br>**(0%)** |
| **9** | **(N=230)** | 94<br>(40.9%) | 59<br>(25.7%) | 15<br>(6.5%) | 9<br>(3.9%) | 50<br>(21.7%) | 3<br>(1.3%) | **0**<br>**(0%)** |
| **10** | **(N=128)** | 102<br>(79.7%) | 5<br>(3.9%) | 2<br>(1.6%) | 2<br>(1.6%) | 16<br>(12.5%) | 1<br>(0.8%) | **0**<br>**(0%)** |
| **11** | **(N=103)** | 80<br>(77.7%) | 5<br>(4.9%) | 2<br>(1.9%) | 1<br>(1%) | 14<br>(13.6%) | 1<br>(1%) | **0**<br>**(0%)** |
| **Mean:** | **(N=2,345)** | **1,052**<br>**(44.9%)** | **553**<br>**(23.6%)** | **135**<br>**(5.8%)** | **89**<br>**(3.8%)** | **482**<br>**(20.6%)** | **34**<br>**(1.4%)** | **0**<br>**(0%)** |

**N** = number of records incorrectly removed by each individual filter. **Categories:** 1. uses human participants or data; 2. review or overview of a condition or intervention affecting humans; 3. review or overview of a factor affecting human health; 4. study outcomes relevant to diagnosing or treating humans; 5. study outcomes relevant to a factor affecting human health; 6. not on animals/nonhumans or human health directly; 7. other. **NB:** Figures in bold represent the lowest numbers and percentages, whereas figures with light grey cells represent the highest numbers and percentages.

Of the seven categories, ‘uses human participants or data’ (category 1) was the most excluded at 44.9% on average. The second most excluded was ‘review or overview of a condition or intervention affecting humans’ (category 2) at 23.6%. This was followed by ‘study outcomes relevant to a factor affecting human health’ (category 5) at 20.6%; ‘review or overview of a factor affecting human health’ (category 3) at 5.8%; ‘study outcomes relevant to diagnosing or treating humans’ (category 4) at 3.8%; ‘not on animals/nonhumans or human health directly’ (category 6) at 1.4%; and the least excluded was ‘other’ at 0%.

Table 6 shows the categories of incorrectly removed records ordered by most to least excluded. The average order is shown and was the same for all filters except methods 10 and 11, which are listed with bold text to highlight their differences from the average. This data should be understood in the context of Table 5, which shows the individual numbers of records incorrectly removed for each filter and category.

**Table 6:** Most to least excluded records by category (All Datasets)

| **Order:**<br>(from most to least excluded) | **Average Order:** | **Filter 10 Order:** | **Filter 11 Order:** |
| --- | --- | --- | --- |
| **First** | Category 1 | Category 1 | Category 1 |
| **Second** | Category 2 | **Category 5** | **Category 5** |
| **Third** | Category 5 | **Category 2** | **Category 2** |
| **Fourth** | Category 3 | Category 3 | Category 3 |
| **Fifth** | Category 4 | Category 4 | **Categories 4 & 6**<br>**(Joint)** |
| **Sixth** | Category 6 | Category 6 |  |
| **Seventh** | Category 7 | Category 7 | Category 7 |

Methods 10 and 11 both differed from the average order by having records with ‘study outcomes relevant to a factor affecting human health’ (category 5) placing as the second most excluded category rather than third, followed by ‘review or overview of a condition or intervention affecting humans’ (category 2) placing third rather than second. The only other difference was for method 11, where records with ‘study outcomes relevant to diagnosing or treating humans’ (category 4) and ‘not on animals/nonhumans or human health directly’ (category 6) were jointly second-to-last.

## DISCUSSION

### How do the filters perform and compare?

Looking across the datasets, the filters with highest sensitivity and specificity (methods 11 and 3, respectively) were the same for Table 3 (humans dataset) and Table 4 (all datasets). The same findings were observed for each dataset individually, which can be seen in the supplementary material. The consistency of this finding is valuable since the datasets included overlapping and irrelevant topics (i.e., both animals/nonhumans and humans, and animals or nonhumans). This suggests that the high sensitivity of method 11 and the high specificity of method 3 in comparison to the other filters, could be generalisable across a broad range of topics.

The filters which achieved the highest overall accuracy differed between the datasets. Method 10 had the highest percentage of correct records (84.8%) for Table 3 (humans dataset), whereas method 3 achieved the highest percentage (73%) for Table 4 (all datasets). This will be due to the different proportions of records in the ‘should be retained’ and ‘should be removed’ categories between the datasets, caused by the different topics that were sought (i.e., animals or nonhumans; humans; and both animals/nonhumans and humans). Although we report on the overall accuracy of the filters, records on animals/nonhumans may be unlikely to make up a significant proportion of results for reviews interested in humans as a subject or population. Therefore, we recommend choosing a filter based on sensitivity rather than overall accuracy. Where specificity is an important consideration, this is best where sensitivity also remains high. As an example, if we compare filters 10 and 11 in Table 4 (all datasets), method 10 is 2.3% less sensitive than method 11, with 8.7% higher specificity.

### Do the different approaches of the filters impact their performance?

A key difference in filter design that affected performance is the subject headings used to remove animal or nonhuman records. Table 1 shows that nine of the 11 methods employed subject headings to remove both animal and nonhuman records, with the remaining two methods (methods 11 and 10) the only ones not to remove nonhuman records. These filters, which were the two most sensitive, are also comparable in their sole use of the ‘animal experiment/’ subject heading, as ‘animal/’ appears in all other methods. Although other variations in subject heading choice can be seen across the filters, these had a much less noticeable impact on performance.

The similarities and differences in the design of the filters is reflected in the sensitivity and specificity they achieved. Although method 11 achieved the highest sensitivity at 88.3% for Table 3 (humans dataset) and 90.6% for Table 4 (all datasets), method 10 was close behind at 86.9% and 88.3%, respectively. All other filters performed similarly in terms of sensitivity, ranging from 75.4% to 78.5%. for Table 3 (humans dataset) and 75.2% to 78.9%. for Table 4 (all datasets). This pattern was also observed for specificity: method 3 achieved highest specificity at 91.7% for Table 3 (humans dataset) and 71.5% for Table 4 (all datasets) but had a similar performance to most other filters. For Table 3 (humans dataset), all other filters ranged from 90.1% to 91.3%, except for methods 11 and 10 where specificity was lowest at 73.4% and 80.4%, respectively. For Table 4 (all datasets), all other filters ranged from 68% to 68.5%, except for methods 11 and 10 at 41.2% and 49.9%, respectively. The distinct differences in the design of methods 11 and 10 compared with the other filters is also reflected in Table 6, as these were the only filters which deviated from the average order of categories that were incorrectly removed.

Another notable difference in filter design that affected performance is the search fields. Table 1 shows that eight of the 11 methods solely employed subject headings; two methods employed the title field in addition to subject headings (methods 4 and 10); and one method employed the title and abstract fields in addition to subject headings (method 3).

Method 3 was the only filter which employed the title and abstract fields (in addition to subject headings) in its approach to remove records. Notably, this filter had highest specificity and lowest sensitivity for both Table 3 (humans dataset) and Table 4 (all datasets). Since terms appearing in these fields may not necessarily reflect the content of a record, their use could have caused records to be incorrectly excluded. Some terms might be used to denote irrelevance: ‘animal studies were excluded’ or might be used metaphorically rather than literally, as Finnegan et al have observed [13]. Since this filter is almost identical to method 4 but unique in its application of the abstract field, it is this aspect of its design which led to it achieving highest specificity and lowest sensitivity.

For the other two filters that employed the title field and subject headings (methods 4 and 10), performance varied. Method 4 ranked sixth for sensitivity, whereas method 10 was the second most sensitive filter overall. Additional key differences between these filters in their use of subject headings (as described above) caused this difference in performance. A further difference is that method 10 removes records with animal terms in terms in the tile where these occur with the ‘animal experiment/’ subject heading. As all 11 filters remove irrelevant free-text terms or subject headings unless they occur with a relevant subject heading, method 10 is unique in its structure as the only filter to remove records with irrelevant free-text terms that occur with an irrelevant subject heading. However, this also means that it is possible for method 10 to remove records with the Emtree heading ‘human/’, in contrast with all other filters. If we compare the similar designs and performance of method 10 and method 11, the unique differences in the structure of method 10 led to its correct removal of more animal records at a slight cost to sensitivity. Notably, there were only minor differences in sensitivity between methods 11 and 10 in Tables 3 and 4 (1.4% and 2.3%, respectively), with larger differences in specificity (7% and 8.7%, respectively).

Information about the origins or usage of the filters may reflect the context for which they were originally developed, which could explain some of the differences in design between the filters. As an example, method 5’s use by the Cochrane Common Mental Disorders group could mean it was developed using records on common mental disorders. Similarly, method 3’s origins as part of the strategy to populate the NHS Economic Evaluations Database could mean it was developed using this study type. This might also explain aspects of its design, such as the use of the title and abstract fields, which may have been valuable for economic evaluations. In the absence of this information, it is useful to examine how the filters compare to each other for the study types used within this project (systematic reviews and RCTs).

As a further point on filter design, several Emtree headings for human study types listed in Elsevier’s 2026 indexing guide are not routinely used in filters that remove animal or nonhuman records [14]. These include: ‘normal human/’, ‘major clinical study/’, ‘clinical article/’, ‘case report/’, ‘human experiment/’, ‘human tissue/’, and ‘human cell/’ [14]. Future studies could investigate if incorporating these subject headings into a filter could increase sensitivity, though it is worth noting that some review teams may not be interested in Emtree terms such as ‘human tissue/’ or ‘human cell/’ where this level of evidence is not relevant. A wider variety of terms for humans are searched for in the free-text fields of the Embase.com search strategy used to populate Cochrane CENTRAL. The animal studies filter for the Embase.com strategy was designed by Julie Glanville and includes terms such as patient, patients, children, man, men, women, woman, pediatric, paediatric, as well as other terms [18]. This suggests the value of including these terms for sensitivity and this approach could also be investigated for Ovid Embase or other databases and platforms.

### How might indexing affect filter performance?

A poster by Soudant and colleagues reported that most records incorrectly excluded by animal filters in Embase were conference abstracts [12]. This finding may suggest that records which only receive automated indexing are more likely to be incorrectly excluded by animal filters. This includes the following types of record: certain non-English language records; pre-prints; conference abstracts; ‘in press’ or ‘in process’ records; records *without* one or more Emtree terms for diseases, drugs, or devices; and records *not identified as*: reviews; short surveys (for case reports only); meta-analyses; and systematic reviews. Information specialists should keep this in mind for review topics meeting these criteria (e.g., those interested in international evidence, conference abstracts, pre-prints, or study types such as economic evaluations). As a result of this finding, Soudant et al discourage the use of filters in strategies aiming to find conference abstracts [12]. However, if information specialists wanted to be cautious about the potential exclusion of these records, they could limit to these records (e.g., conference abstracts or pre-prints) separately in the search strategy and then use the OR operator to pool these records with results applying an animal/nonhuman filter.

As records categorised with ‘uses human participants or data’ (category 1) were the most incorrectly excluded, this suggests records may not always indexed as we might expect. This warrants further investigation to understand if this occurs for certain types of records only (i.e, certain study types, publication types, or for records with automated indexing only).

### What do we mean by an animal or nonhuman record?

In evidence syntheses, definitions of animal/nonhuman records that are ineligible and should be excluded (both in the searches and during screening) could vary depending on the specific review being conducted. This suggests the need for discussions between information specialists and review teams about the types of animal or nonhuman studies they are aiming to exclude, and whether animals or nonhumans could feature in relevant studies. To provide examples, a review interested in assessing all interventions for mental health conditions could miss records on animal-assisted therapies, or reviews interested in methods to detect cancer could miss records on detection dogs, where a limit to remove animals/nonhumans was applied and where Emtree terms such as ‘human/’, had not been applied to the record.

## LIMITATIONS

This is an exploratory study in which only 2,689 records in Embase taken from healthcare literature were tested. As records were retrieved using filters for systematic reviews and RCTs, filter performance could vary for other study types. As we did not intend to test differences in filter performance by study type, we did not use export the same number of records for each study type. For all datasets, only titles and abstracts were double screened.

Screening decisions about what is or is not an ‘animal’ or ‘nonhuman’ record were subjective, and definitions could vary. For these reasons, a broad approach was taken for greater generalisability of the findings.

Categorisation of the content or focus of removed records that ‘should be retained’ was performed by one reviewer only based on the titles and abstracts.

## CONCLUSION

Between information specialists, review teams, and even on a review-by-review basis, there may be different definitions or criteria for excluding animal or nonhuman records. Information specialists and review teams should discuss this in the context of the specific condition, phenomenon of interest, intervention, or level of evidence needed for the review. Detailed understanding of a review team’s inclusion and exclusion criteria helps ensure potentially eligible records are not removed through filter use.

Where filter use is considered appropriate to remove animal or nonhuman records in Ovid Embase, we recommend the use of method 11 due to its sensitivity. Our investigation of how Emtree terms are indexed suggests that performance of the filters could vary by subject, publication type, study type, and language, which information specialists should keep in mind. Further research could be conducted to test how the filters perform for conference abstracts, pre-prints, non-English language records and subjects that are unlikely to have one or more Emtree terms for diseases, drugs, or devices. Further testing and validation of the filters for different study types, such as qualitative studies or economic evaluations, would also be useful.

Overall, our findings are informative to help information specialists understand the impact of 11 different methods in Ovid Embase to remove animal or nonhuman records, which are compared in terms of both performance and design.

## Supporting information

Appendix - Search Strategies

## ACKNOWLEDGEMENTS

We would like to acknowledge Janet Clapton for commenting on an earlier draft of this paper and Ovid’s Customer Success Team and Elsevier for their time and help answering our enquiries.

## ETHICS STATEMENT

Ethical approval was not necessary for this study.

## PEER REVIEW STATEMENT

This study was originally submitted to a journal in December 2024 and received one round of peer reviews. The authors decided to withdraw the study in February 2026 and submit to another journal. All data was checked and was up to date at the time of submission to medRxiv.

## DATA AVAILABILITY STATEMENT

Data associated with this article are available in the appendix and supplementary material or can be obtained by contacting the first author.

