## Appendix - Search Strategies for "Removing animal and nonhuman records in Ovid Embase: A comparison of 11 filters"

### **Dataset – Animals/Nonhumans:**

#### **Lines to Retrieve Records for Animals/Nonhumans Dataset**

Embase via Ovid http://ovidsp.ovid.com/

Database Segment and Indexing: <1974 to 2024 June 28>

Date Searched: 1st July 2024

1	animal welfare.af. (34205)

2	veterinary parasitology.af. (15999)

3	veterinary surgery.af. (7413)

4	veterinary dentistry.af. (304)

5	animal dentistry.af. (57)

6	equine dentistry.af. (108)

7	aflatoxicosis.af. (805)

8	bluetongue.af. (3862)

9	cattle tick.af. (1017)

10	coccidiosis.af. (7391)

11	leptospirosis.af. (11830)

12	neosporosis.af. (1426)

13	wooden tongue.af. (57)

14	shellfish immunology.af. (327)

15	dutch elm disease.af. (141)

16	ophiocordyceps unilateralis.af. (27)

17	or/1-16 (82989)

18	systematic$ review$.ti,ab. (410334)

19	systematic$ literature review$.ti,ab. (28631)

20	"systematic review"/ (473545)

21	"systematic review (topic)"/ (35217)

22	meta analysis/ (320342)

23	"meta analysis (topic)"/ (56278)

24	meta-analytic$.ti,ab. (12292)

25	meta-analysis.ti,ab. (333845)

26	metanalysis.ti,ab. (1485)

27	metaanalysis.ti,ab. (10710)

28	meta analysis.ti,ab. (333845)

29	meta-synthesis.ti,ab. (1904)

30	metasynthesis.ti,ab. (569)

31	meta synthesis.ti,ab. (1904)

32	meta-regression.ti,ab. (17851)

33	metaregression.ti,ab. (1589)

34	meta regression.ti,ab. (17851)

35	(synthes$ adj3 literature).ti,ab. (7512)

36	(synthes$ adj3 evidence).ti,ab. (22111)

37	(synthes$ adj2 qualitative).ti,ab. (7555)

38	integrative review.ti,ab. (5422)

39	data synthesis.ti,ab. (17753)

40	(research synthesis or narrative synthesis).ti,ab. (9265)

41	(systematic study or systematic studies).ti,ab. (17095)

42	(systematic comparison$ or systematic overview$).ti,ab. (5552)

43	(systematic adj2 search$).ti,ab. (62464)

44	systematic$ literature research$.ti,ab. (543)

45	(review adj3 scientific literature).ti,ab. (2897)

46	(literature review adj2 side effect$).ti,ab. (24)

47	(literature review adj2 adverse effect$).ti,ab. (10)

48	(literature review adj2 adverse event$).ti,ab. (21)

49	(evidence-based adj2 review).ti,ab. (4969)

50	comprehensive review.ti,ab. (33841)

51	critical review.ti,ab. (21992)

52	critical analysis.ti,ab. (10761)

53	quantitative review.ti,ab. (1007)

54	structured review.ti,ab. (1455)

55	realist review.ti,ab. (757)

56	realist synthesis.ti,ab. (402)

57	(pooled adj2 analysis).ti,ab. (33479)

58	(pooled data adj6 (studies or trials)).ti,ab. (4316)

59	(medline and (inclusion adj3 criteria)).ti,ab. (41638)

60	(search adj (strateg$ or term$)).ti,ab. (60127)

61	or/18-60 (945654)

62	medline.ab. (222986)

63	pubmed.ab. (296450)

64	cochrane.ab. (190840)

65	embase.ab. (217402)

66	cinahl.ab. (58572)

67	psyc?lit.ab. (1012)

68	psyc?info.ab. (62148)

69	lilacs.ab. (12574)

70	(literature adj3 search$).ab. (128249)

71	(database$ adj3 search$).ab. (134000)

72	(bibliographic adj3 search$).ab. (4961)

73	(electronic adj3 search$).ab. (44060)

74	(electronic adj3 database$).ab. (68208)

75	(computeri?ed adj3 search$).ab. (5026)

76	(internet adj3 search$).ab. (6095)

77	included studies.ab. (62285)

78	(inclusion adj3 studies).ab. (34855)

79	inclusion criteria.ab. (243967)

80	selection criteria.ab. (47347)

81	predefined criteria.ab. (3712)

82	predetermined criteria.ab. (1718)

83	(assess$ adj3 (quality or validity)).ab. (163525)

84	(select$ adj3 (study or studies)).ab. (128134)

85	(data adj3 extract$).ab. (148505)

86	extracted data.ab. (30123)

87	(data adj2 abstracted).ab. (12241)

88	(data adj3 abstraction).ab. (3736)

89	published intervention$.ab. (331)

90	((study or studies) adj2 evaluat$).ab. (383830)

91	(intervention$ adj2 evaluat$).ab. (23090)

92	confidence interval$.ab. (739122)

93	heterogeneity.ab. (331257)

94	pooled.ab. (195113)

95	pooling.ab. (23675)

96	odds ratio$.ab. (500130)

97	(Jadad or coding).ab. (289769)

98	evidence-based.ti,ab. (213603)

99	or/62-98 (3028500)

100	review.pt. (3243148)

101	99 and 100 (421773)

102	review.ti. (867209)

103	99 and 102 (370138)

104	(review$ adj10 (papers or trials or trial data or studies or evidence or intervention$ or evaluation$ or outcome$ or findings)).ti,ab. (824317)

105	(retriev$ adj10 (papers or trials or studies or evidence or intervention$ or evaluation$ or outcome$ or findings)).ti,ab. (46542)

106	61 or 101 or 103 or 104 or 105 (1599909)

107	letter.pt. (1327839)

108	editorial.pt. (810830)

109	107 or 108 (2138669)

110	106 not 109 (1568161)

111	randomized controlled trial/ (828898)

112	controlled clinical trial/ (473459)

113	Random$.ti,ab,ot. (2085562)

114	randomization/ (99489)

115	intermethod comparison/ (307279)

116	placebo.ti,ab,ot. (379291)

117	(compare or compared or comparison).ti,ot. (628450)

118	((evaluated or evaluate or evaluating or assessed or assess) and (compare or compared or comparing or comparison)).ab. (2948399)

119	(open adj label).ti,ab,ot. (116367)

120	((double or single or doubly or singly) adj (blind or blinded or blindly)).ti,ab,ot. (283916)

121	double blind procedure/ (220584)

122	parallel group$1.ti,ab,ot. (33780)

123	(crossover or cross over).ti,ab,ot. (128969)

124	((assign$ or match or matched or allocation) adj5 (alternate or group or groups or intervention or interventions or patient or patients or subject or subjects or participant or participants)).ti,ab,ot. (436164)

125	(assigned or allocated).ti,ab,ot. (515550)

126	(controlled adj7 (study or design or trial)).ti,ab,ot. (475422)

127	(volunteer or volunteers).ti,ab,ot. (290432)

128	trial.ti,ot. (428134)

129	or/111-128 (6183681)

130	110 or 129 (7290857)

131	17 (82989)

132	130 and 131 (14717)

133	132 (14717)

134	(animal/ or animal experiment/ or animal model/ or animal tissue/ or nonhuman/) not exp human/ (7017940)

135	133 not 134 (3298)

136	133 and 134 (11419)

137	(animal/ or nonhuman/) not exp human/ (6676639)

138	133 not 137 (3330)

139	133 and 137 (11387)

140	animal/ or exp animal experiment/ or nonhuman/ (9784451)

141	(rat or rats or mouse or mice or hamster or hamsters or animal or animals or dog or dogs or cat or cats or bovine or sheep).ti,sh. (5707457)

142	or/140-141 (10345333)

143	exp human/ or human experiment/ (26722690)

144	142 not (142 and 143) (7385781)

145	133 not 144 (3138)

146	133 and 144 (11579)

147	animal/ or exp animal experiment/ or nonhuman/ (9784451)

148	(rat or rats or mouse or mice or hamster or hamsters or animal or animals or dog or dogs or cat or cats or bovine or sheep).ti,ab,sh. (6681690)

149	or/147-148 (10749553)

150	exp human/ or human experiment/ (26722690)

151	149 not (149 and 150) (7521649)

152	133 not 151 (3024)

153	133 and 151 (11693)

154	((animal or nonhuman) not human).sh. (6678852)

155	133 not 154 (3329)

156	133 and 154 (11388)

157	animal experiment/ not (human experiment/ or human/) (2644485)

158	133 not 157 (10088)

159	133 and 157 (4629)

160	(exp animal/ or exp invertebrate/ or nonhuman/ or animal experiment/ or animal tissue/ or animal model/ or exp plant/ or exp fungus/) not (exp human/ or human tissue/) (7956295)

161	133 not 160 (3113)

162	133 and 160 (11604)

163	(exp animal/ or exp invertebrate/ or nonhuman/ or animal experiment/ or animal tissue/ or animal model/ or exp plant/ or exp fungus/) not (exp human/ or human tissue/ or human experiment/) (7955132)

164	133 not 163 (3124)

165	133 and 163 (11593)

166	(exp animal/ or exp invertebrate/ or nonhuman/ or animal experiment/ or animal tissue/ or animal model/) not (exp human/ or human tissue/ or human experiment/) (7467817)

167	133 not 166 (3127)

168	133 and 166 (11590)

169	(exp animals/ or exp animal experimentation/ or exp animal experiment/ or exp models animal/ or nonhuman/ or exp vertebrate/ or exp vertebrates/) not (exp humans/ or exp human experimentation/ or exp human experiment/) (7469930)

170	133 not 169 (3101)

171	133 and 169 (11616)

172	(rat or rats or mouse or mice or swine or porcine or murine or sheep or lambs or pigs or piglets or rabbit or rabbits or cat or cats or dog or dogs or cattle or bovine or monkey or monkeys or trout or marmoset$1).ti. and animal experiment/ (1257365)

173	animal experiment/ not (human experiment/ or human/) (2644485)

174	or/172-173 (2719082)

175	133 not 174 (9902)

176	133 and 174 (4815)

177	136 or 139 or 146 or 153 or 156 or 159 or 162 or 165 or 168 or 171 or 176 (11933)

178	135 or 138 or 145 or 152 or 155 or 158 or 161 or 164 or 167 or 170 or 175 (10113)

**Should be Removed (Animals)**

Embase via Ovid http://ovidsp.ovid.com/

Database Segment and Indexing: <1974 to 2024 November 19>

Date Searched: 20^th^ November 2024

1	("2032969261" or "2032907307" or "2032876113" or "2032866216" or "2032863547" or "2032861218" or "2032861184" or "2032844752" or "2032844544" or "2032771397" or "2032717894" or "2032717891" or "2032717890" or "2032717703" or "2032704664" or "2032688769" or "2032613357" or "2032607695" or "2032598488" or "2032594881" or "2032583002" or "2032520776" or "2032508349" or "2032508265" or "2032497017" or "2032491725" or "2032467286" or "2032460556" or "2032441777" or "2032435764" or "2032434332" or "2032428621" or "2032414157" or "2032408100" or "2032380338" or "2032370322" or "2032313547" or "2032228163" or "2032212892" or "2032149951" or "2032145346" or "2032145326" or "2032027895" or "2032019395" or "2031983172" or "2031970865" or "2031931685" or "2031924149" or "2031922460" or "2031919973" or "2031887039" or "2031825160" or "2031823861" or "2031811625" or "2031773743" or "2031773641" or "2031773403" or "2031768762" or "2031741486" or "2031731696" or "2031712623" or "2031709628" or "2031688618" or "2031683888" or "2031683862" or "2031649872" or "2031648358" or "2031597511" or "2031596762" or "2031596742" or "2031552116" or "2031477758" or "2031477296" or "2031471636" or "2031467776" or "2031461765" or "2031452971" or "2031419717" or "2031416182" or "2031369906" or "2031366093" or "2031343213" or "2031334762" or "2031324242" or "2031301674" or "2031291554" or "2031273341" or "2031263799" or "2031263783" or "2031220708" or "2031182489" or "2031175755" or "2031090821" or "2031078994" or "2031061714" or "2030968615" or "2030947714" or "2030947543" or "2030942861" or "2030895983" or "2030863550" or "2030853291" or "2030760808" or "2030755323" or "2030753865" or "2030749972" or "2030670081" or "2030624266" or "2030608583" or "2030598826" or "2030595936" or "2030508380" or "2030508376" or "2030472497" or "2030418520" or "2030414723" or "2030368806" or "2030344792" or "2030332376" or "2030332323" or "2030318378" or "2030297746" or "2030271097" or "2030270056" or "2030262469" or "2030252717" or "2030250440" or "2030235297" or "2030234009" or "2030229271" or "2030225891" or "2030207108" or "2030196385" or "2030189387" or "2030187495" or "2030187482" or "2030187461" or "2030187416" or "2030187395" or "2030177889" or "2030177764" or "2030173447" or "2030172379" or "2030170506" or "2030129034" or "2030128860" or "2030122690" or "2030106842" or "2030100530" or "2030086481" or "2030075104" or "2030074676" or "2030074542" or "2030043176" or "2030039199" or "2030028798" or "2030020589" or "2030016330" or "2030013604" or "2030000119" or "2029991488" or "2029980292" or "2029980268" or "2029980261" or "2029949521" or "2029939630" or "2029934142" or "2029933778" or "2029933541" or "2029924152" or "2029924149" or "2029910290" or "2029901673" or "2029885798" or "2029885767" or "2029885754" or "2029858088" or "2029844139" or "2029839131" or "2029838488" or "2029838478" or "2029836108" or "2029828076" or "2029813205" or "2029813127" or "2029806348" or "2029791042" or "2029781495" or "2029700358" or "2029694992" or "2029694279" or "2029664010" or "2029645546" or "2029635630" or "2029629545" or "2029606565" or "2029588276" or "2029588252" or "2029584319" or "2029571300" or "2029544887" or "2029540291" or "2029528416" or "2029494804" or "2029487318" or "2029481298" or "2029473654" or "2029464808" or "2029456015" or "2029454737" or "2029426534" or "2029422429" or "2029396580" or "2029386665" or "2029386663" or "2029386655" or "2029384238" or "2029384224" or "2029384216" or "2029384184" or "2029351876" or "2029305113" or "2029295035" or "2029263336" or "2029263181" or "2029259571" or "2029255180" or "2029243985" or "2029243964" or "2029215167" or "2029214352" or "2029202899" or "2029194124" or "2029190494" or "2029190466" or "2029184158" or "2029183888" or "2029177117" or "2029175136" or "2029171578" or "2029152965" or "2029152964" or "2029152957" or "2029152944" or "2029152934" or "2029152932" or "2029152920" or "2029149423" or "2029149402" or "2029149361" or "2029149339" or "2029148958" or "2029137244" or "2029135399" or "2029132903" or "2029123764" or "2029080519" or "2029075693" or "2029075592" or "2029073213" or "2029045450" or "2028980109" or "2028964196" or "2028964070" or "2028963577" or "2028954059" or "2028933611" or "2028931705" or "2028886161" or "2028880179" or "2028865492" or "2028864649" or "2028830101" or "2028827334" or "2028770753" or "2028745522" or "2028735686" or "2028730490" or "2028707744" or "2028663467" or "2028662277" or "2028660767" or "2028660734" or "2028659968" or "2028659943" or "2028659929" or "2028659926" or "2028629558" or "2028629463" or "2028600070" or "2028597928" or "2028527647" or "2028523451" or "2028520915" or "2028520911" or "2028520888" or "2028520883" or "2028520845" or "2028458347" or "2028427778" or "2028378120" or "2028342035" or "2028296577" or "2028288410" or "2028261223" or "2028261212" or "2028211716" or "2028181872" or "2028179599" or "2028175624" or "2028153254" or "2028141454" or "2028133421" or "2028133417" or "2028106791" or "2028088041" or "2028087965" or "2028076134" or "2028068358" or "2028068241" or "2028068162" or "2028041500" or "2028035378" or "2028011417" or "2027969386" or "2027967113" or "2027963316" or "2027962180" or "2027952637" or "2027947372" or "2027932074" or "2027898614" or "2027848001" or "2027847730" or "2027639492" or "2027322935" or "2027319659" or "2027303298" or "2027301883" or "2027301881" or "2027179972" or "2027178341" or "2027073137" or "2027048669" or "2027001492" or "2026987366" or "2026943498" or "2026908199" or "2026749002" or "2026533335" or "2026450084" or "2026425458" or "2026424961" or "2026319925" or "2026234920" or "2026222069" or "2026118401" or "2025995005" or "2025978809" or "2025953403" or "2025952617" or "2025947798" or "2025882341" or "2025851152" or "2025828102" or "2025675256" or "2025629290" or "2025619865" or "2025613834" or "2025411769" or "2025312416" or "2025312110" or "2025173216" or "2025014415" or "2025001955" or "2025001478" or "2024928360" or "2024860440" or "2024845330" or "2024539355" or "2024515879" or "2024459407" or "2024397976" or "2024392990" or "2024392989" or "2024164332" or "2023944912" or "2023663201" or "2023661743" or "2023541818" or "2023539953" or "2023538167" or "2022257449" or "2022244019" or "2022241968" or "2022131152" or "2022025322" or "2021965350" or "2021572889" or "2021263154" or "2020888008" or "2020468685" or "2019769272" or "2019455970" or "2019187514" or "2018968536" or "2018791891" or "2018442273" or "2018286492" or "2018200099" or "2017208568" or "2016822203" or "2016437827" or "2016436531" or "2016149918" or "2016019686" or "2015954495" or "2015896251" or "2015612779" or "2015568501" or "2015543334" or "2015408604" or "2014716567" or "2014004473" or "2013543428" or "2013520450" or "2007786379" or "2005540458" or "2005489255" or "1369919611" or "1369919610" or "1369919608" or "1369168304" or "1364799797" or "644584608" or "644530544" or "644514997" or "644514954" or "644510248" or "644476050" or "644462972" or "644455169" or "644414666" or "644397159" or "644396275" or "644383145" or "644360986" or "644352410" or "644349209" or "644333717" or "644333157" or "644333148" or "644288033" or "644277419" or "644266897" or "644266510" or "644219942" or "644202936" or "644168532" or "644127616" or "644112451" or "644083935" or "644076054" or "644046671" or "644046604" or "644037741" or "644026777" or "644023786" or "644016118" or "644013672" or "643998069" or "643997863" or "643974588" or "643974324" or "643972967" or "643945226" or "643899613" or "643899582" or "643896493" or "643804693" or "643771267" or "643770636" or "643767720" or "643765963" or "643750449" or "643743888" or "643705325" or "643691538" or "643642788" or "643642778" or "643634401" or "643627904" or "643624893" or "643607894" or "643606453" or "643584771" or "643572522" or "643572078" or "643554290" or "643553795" or "643553214" or "643531586" or "643527368" or "643523453" or "643518601" or "643501595" or "643494950" or "643491495" or "643476733" or "643473989" or "643456782" or "643407734" or "643378924" or "643347205" or "643320297" or "643303458" or "643286504" or "643281122" or "643239812" or "643211618" or "643199766" or "643175350" or "643172880" or "643159850" or "643159582" or "643149277" or "643088149" or "643087724" or "643071713" or "643062931" or "643054679" or "643035597" or "643033993" or "642902715" or "642889446" or "642877426" or "642854570" or "642790163" or "642789303" or "642718219" or "642715173" or "642691083" or "642603242" or "642568960" or "642523603" or "642277742" or "642277698" or "642277329" or "642218572" or "642218454" or "642218370" or "642217760" or "642217585" or "642217531" or "642138460" or "642138023" or "642137991" or "641867420" or "641754483" or "641488118" or "641323556" or "640017437" or "639854301" or "639462718" or "639462687" or "639462666" or "639462638" or "639462636" or "639462601" or "639462587" or "639462569" or "639462423" or "639462396" or "639462331" or "639462314" or "639462296" or "639344629" or "639344589" or "639344442" or "639178996" or "638853410" or "638853403" or "638844738" or "638844731" or "638844728" or "638844713" or "638844704" or "638844700" or "638844697" or "638696213" or "638696210" or "638696199" or "638696192" or "638696187" or "638696049" or "638696032" or "638696019" or "638549716" or "637909231" or "637397557" or "637321535" or "636976096" or "636949046" or "635720901" or "635573009" or "635235363" or "635143167" or "634981853" or "634453348" or "634029889" or "633950189" or "633401499" or "633191042" or "631512973" or "631108772" or "630424482" or "630403676" or "629537457" or "629356275" or "628951272" or "627182976" or "626822374" or "626101980" or "624713952" or "620636160" or "613811929" or "50733091").an. (623)

2	remove duplicates from 1 (623)

3	(animal/ or animal experiment/ or animal model/ or animal tissue/ or nonhuman/) not exp human/ (7127952)

4	2 not 3 (157)

5	2 and 3 (466)

6	(animal/ or nonhuman/) not exp human/ (6786390)

7	2 not 6 (157)

8	2 and 6 (466)

9	animal/ or exp animal experiment/ or nonhuman/ (9967558)

10	(rat or rats or mouse or mice or hamster or hamsters or animal or animals or dog or dogs or cat or cats or bovine or sheep).ti,sh. (5792466)

11	or/9-10 (10530178)

12	exp human/ or human experiment/ (27323434)

13	11 not (11 and 12) (7496062)

14	2 not 13 (154)

15	2 and 13 (469)

16	animal/ or exp animal experiment/ or nonhuman/ (9967558)

17	(rat or rats or mouse or mice or hamster or hamsters or animal or animals or dog or dogs or cat or cats or bovine or sheep).ti,ab,sh. (6782932)

18	or/16-17 (10939926)

19	exp human/ or human experiment/ (27323434)

20	18 not (18 and 19) (7633032)

21	2 not 20 (146)

22	2 and 20 (477)

23	((animal or nonhuman) not human).sh. (6788704)

24	2 not 23 (157)

25	2 and 23 (466)

26	animal experiment/ not (human experiment/ or human/) (2685603)

27	2 not 26 (437)

28	2 and 26 (186)

29	(exp animal/ or exp invertebrate/ or nonhuman/ or animal experiment/ or animal tissue/ or animal model/ or exp plant/ or exp fungus/) not (exp human/ or human tissue/) (8076067)

30	2 not 29 (151)

31	2 and 29 (472)

32	(exp animal/ or exp invertebrate/ or nonhuman/ or animal experiment/ or animal tissue/ or animal model/ or exp plant/ or exp fungus/) not (exp human/ or human tissue/ or human experiment/) (8074833)

33	2 not 32 (151)

34	2 and 32 (472)

35	(exp animal/ or exp invertebrate/ or nonhuman/ or animal experiment/ or animal tissue/ or animal model/) not (exp human/ or human tissue/ or human experiment/) (7578504)

36	2 not 35 (151)

37	2 and 35 (472)

38	(exp animals/ or exp animal experimentation/ or exp animal experiment/ or exp models animal/ or nonhuman/ or exp vertebrate/ or exp vertebrates/) not (exp humans/ or exp human experimentation/ or exp human experiment/) (7580901)

39	2 not 38 (151)

40	2 and 38 (472)

41	(rat or rats or mouse or mice or swine or porcine or murine or sheep or lambs or pigs or piglets or rabbit or rabbits or cat or cats or dog or dogs or cattle or bovine or monkey or monkeys or trout or marmoset$1).ti. and animal experiment/ (1275917)

42	animal experiment/ not (human experiment/ or human/) (2685603)

43	or/41-42 (2763141)

44	2 not 43 (364)

45	2 and 43 (259)

46	5 (466)

47	8 (466)

48	15 (469)

49	22 (477)

50	25 (466)

51	28 (186)

52	31 (472)

53	34 (472)

54	37 (472)

55	40 (472)

56	45 (259)

**Should be Retained (Not Animals)**

Embase via Ovid http://ovidsp.ovid.com/

Database Segment and Indexing: <1974 to 2024 November 19>

Date Searched: 20^th^ November 2024

Two accession numbers (i.e., two records) were removed from this dataset prior to testing where there was uncertainty about whether the record should be retained.

1	("2034394337" or "2032950631" or "2032909485" or "2032771772" or "2032720124" or "2032720123" or "2032680034" or "2032660776" or "2032608001" or "2032435246" or "2032382905" or "2032283215" or "2032213028" or "2032150106" or "2031826923" or "2031823453" or "2031752330" or "2031632352" or "2031506511" or "2031470894" or "2031394826" or "2031386474" or "2031292174" or "2031183114" or "2031175758" or "2031170465" or "2031135945" or "2031089068" or "2031058328" or "2031006418" or "2031005101" or "2031002182" or "2030826032" or "2030727482" or "2030521505" or "2030397400" or "2030322494" or "2030235106" or "2030221449" or "2030212203" or "2030181271" or "2030047923" or "2030038251" or "2030007139" or "2029965494" or "2029924182" or "2029879726" or "2029866880" or "2029857886" or "2029800773" or "2029741316" or "2029653041" or "2029646460" or "2029635567" or "2029635162" or "2029565031" or "2029490307" or "2029448005" or "2029442073" or "2029441471" or "2029422012" or "2029421830" or "2029407880" or "2029361295" or "2029347975" or "2029258990" or "2029211348" or "2029196909" or "2029175727" or "2029175726" or "2029149387" or "2029064315" or "2029059180" or "2029058279" or "2028997854" or "2028965509" or "2028825860" or "2028803323" or "2028593107" or "2028503298" or "2028494494" or "2028423884" or "2028261002" or "2028194311" or "2028130533" or "2028075455" or "2028036998" or "2027993188" or "2027972980" or "2027944607" or "2027849811" or "2027848981" or "2027848002" or "2027785892" or "2027772510" or "2027734972" or "2027734157" or "2027245256" or "2027192276" or "2026890439" or "2026822821" or "2026811161" or "2026469789" or "2026290018" or "2025856530" or "2025689373" or "2025173222" or "2024755348" or "2024174492" or "2022870539" or "2022074225" or "2021629015" or "2020755740" or "2019770967" or "2019455869" or "2017754218" or "2017158748" or "2015874265" or "2014062030" or "2010048104" or "2007786479" or "2004291033" or "2001672726" or "644589887" or "644561863" or "644534549" or "644519809" or "644510056" or "644485422" or "644461355" or "644447459" or "644334816" or "644256000" or "644056962" or "643997978" or "643760086" or "643760085" or "643567614" or "643566330" or "643564439" or "643508604" or "643071756" or "642818617" or "641654494" or "638826889" or "637302637" or "636632816" or "636478876" or "634932623" or "634930622" or "633306989" or "632053853" or "631271087" or "624263703" or "362270316").an. (151)

2	remove duplicates from 1 (151)

3	(animal/ or animal experiment/ or animal model/ or animal tissue/ or nonhuman/) not exp human/ (7127952)

4	2 not 3 (128)

5	2 and 3 (23)

6	(animal/ or nonhuman/) not exp human/ (6786390)

7	2 not 6 (128)

8	2 and 6 (23)

9	animal/ or exp animal experiment/ or nonhuman/ (9967558)

10	(rat or rats or mouse or mice or hamster or hamsters or animal or animals or dog or dogs or cat or cats or bovine or sheep).ti,sh. (5792466)

11	or/9-10 (10530178)

12	exp human/ or human experiment/ (27323434)

13	11 not (11 and 12) (7496062)

14	2 not 13 (128)

15	2 and 13 (23)

16	animal/ or exp animal experiment/ or nonhuman/ (9967558)

17	(rat or rats or mouse or mice or hamster or hamsters or animal or animals or dog or dogs or cat or cats or bovine or sheep).ti,ab,sh. (6782932)

18	or/16-17 (10939926)

19	exp human/ or human experiment/ (27323434)

20	18 not (18 and 19) (7633032)

21	2 not 20 (120)

22	2 and 20 (31)

23	((animal or nonhuman) not human).sh. (6788704)

24	2 not 23 (128)

25	2 and 23 (23)

26	animal experiment/ not (human experiment/ or human/) (2685603)

27	2 not 26 (143)

28	2 and 26 (8)

29	(exp animal/ or exp invertebrate/ or nonhuman/ or animal experiment/ or animal tissue/ or animal model/ or exp plant/ or exp fungus/) not (exp human/ or human tissue/) (8076067)

30	2 not 29 (127)

31	2 and 29 (24)

32	(exp animal/ or exp invertebrate/ or nonhuman/ or animal experiment/ or animal tissue/ or animal model/ or exp plant/ or exp fungus/) not (exp human/ or human tissue/ or human experiment/) (8074833)

33	2 not 32 (127)

34	2 and 32 (24)

35	(exp animal/ or exp invertebrate/ or nonhuman/ or animal experiment/ or animal tissue/ or animal model/) not (exp human/ or human tissue/ or human experiment/) (7578504)

36	2 not 35 (127)

37	2 and 35 (24)

38	(exp animals/ or exp animal experimentation/ or exp animal experiment/ or exp models animal/ or nonhuman/ or exp vertebrate/ or exp vertebrates/) not (exp humans/ or exp human experimentation/ or exp human experiment/) (7580901)

39	2 not 38 (127)

40	2 and 38 (24)

41	(rat or rats or mouse or mice or swine or porcine or murine or sheep or lambs or pigs or piglets or rabbit or rabbits or cat or cats or dog or dogs or cattle or bovine or monkey or monkeys or trout or marmoset$1).ti. and animal experiment/ (1275917)

42	animal experiment/ not (human experiment/ or human/) (2685603)

43	or/41-42 (2763141)

44	2 not 43 (131)

45	2 and 43 (20)

46	4 (128)

47	7 (128)

48	14 (128)

49	21 (120)

50	24 (128)

51	27 (143)

52	30 (127)

53	33 (127)

54	36 (127)

55	39 (127)

56	44 (131)

### **Dataset – Humans:**

**Lines to Retrieve Records for Humans Dataset**

Embase via Ovid http://ovidsp.ovid.com/

Database Segment and Indexing: <1974 to 2024 June 28>

Date Searched: 27^th^ June 2024

1	personality disorder*.af. (60157)

2	anger management.af. (848)

3	orthorexia.af. (588)

4	migraine*.af. (91770)

5	rhytidoplasty.af. (4679)

6	genioplasty.af. (1447)

7	mentoplasty.af. (116)

8	wheelchair*.af. (17872)

9	walking frame.af. (185)

10	ethnicity.af. (200540)

11	speech therap*.af. (21209)

12	medical clowning.af. (79)

13	school meal*.af. (1560)

14	adverse childhood experience*.af. (6439)

15	medical cannabis.af. (5254)

16	athlete's foot.af. (321)

17	or/1-16 (409988)

18	systematic$ review$.ti,ab. (410334)

19	systematic$ literature review$.ti,ab. (28631)

20	"systematic review"/ (473545)

21	"systematic review (topic)"/ (35217)

22	meta analysis/ (320342)

23	"meta analysis (topic)"/ (56278)

24	meta-analytic$.ti,ab. (12292)

25	meta-analysis.ti,ab. (333845)

26	metanalysis.ti,ab. (1485)

27	metaanalysis.ti,ab. (10710)

28	meta analysis.ti,ab. (333845)

29	meta-synthesis.ti,ab. (1904)

30	metasynthesis.ti,ab. (569)

31	meta synthesis.ti,ab. (1904)

32	meta-regression.ti,ab. (17851)

33	metaregression.ti,ab. (1589)

34	meta regression.ti,ab. (17851)

35	(synthes$ adj3 literature).ti,ab. (7512)

36	(synthes$ adj3 evidence).ti,ab. (22111)

37	(synthes$ adj2 qualitative).ti,ab. (7555)

38	integrative review.ti,ab. (5422)

39	data synthesis.ti,ab. (17753)

40	(research synthesis or narrative synthesis).ti,ab. (9265)

41	(systematic study or systematic studies).ti,ab. (17095)

42	(systematic comparison$ or systematic overview$).ti,ab. (5552)

43	(systematic adj2 search$).ti,ab. (62464)

44	systematic$ literature research$.ti,ab. (543)

45	(review adj3 scientific literature).ti,ab. (2897)

46	(literature review adj2 side effect$).ti,ab. (24)

47	(literature review adj2 adverse effect$).ti,ab. (10)

48	(literature review adj2 adverse event$).ti,ab. (21)

49	(evidence-based adj2 review).ti,ab. (4969)

50	comprehensive review.ti,ab. (33841)

51	critical review.ti,ab. (21992)

52	critical analysis.ti,ab. (10761)

53	quantitative review.ti,ab. (1007)

54	structured review.ti,ab. (1455)

55	realist review.ti,ab. (757)

56	realist synthesis.ti,ab. (402)

57	(pooled adj2 analysis).ti,ab. (33479)

58	(pooled data adj6 (studies or trials)).ti,ab. (4316)

59	(medline and (inclusion adj3 criteria)).ti,ab. (41638)

60	(search adj (strateg$ or term$)).ti,ab. (60127)

61	or/18-60 (945654)

62	medline.ab. (222986)

63	pubmed.ab. (296450)

64	cochrane.ab. (190840)

65	embase.ab. (217402)

66	cinahl.ab. (58572)

67	psyc?lit.ab. (1012)

68	psyc?info.ab. (62148)

69	lilacs.ab. (12574)

70	(literature adj3 search$).ab. (128249)

71	(database$ adj3 search$).ab. (134000)

72	(bibliographic adj3 search$).ab. (4961)

73	(electronic adj3 search$).ab. (44060)

74	(electronic adj3 database$).ab. (68208)

75	(computeri?ed adj3 search$).ab. (5026)

76	(internet adj3 search$).ab. (6095)

77	included studies.ab. (62285)

78	(inclusion adj3 studies).ab. (34855)

79	inclusion criteria.ab. (243967)

80	selection criteria.ab. (47347)

81	predefined criteria.ab. (3712)

82	predetermined criteria.ab. (1718)

83	(assess$ adj3 (quality or validity)).ab. (163525)

84	(select$ adj3 (study or studies)).ab. (128134)

85	(data adj3 extract$).ab. (148505)

86	extracted data.ab. (30123)

87	(data adj2 abstracted).ab. (12241)

88	(data adj3 abstraction).ab. (3736)

89	published intervention$.ab. (331)

90	((study or studies) adj2 evaluat$).ab. (383830)

91	(intervention$ adj2 evaluat$).ab. (23090)

92	confidence interval$.ab. (739122)

93	heterogeneity.ab. (331257)

94	pooled.ab. (195113)

95	pooling.ab. (23675)

96	odds ratio$.ab. (500130)

97	(Jadad or coding).ab. (289769)

98	evidence-based.ti,ab. (213603)

99	or/62-98 (3028500)

100	review.pt. (3243148)

101	99 and 100 (421773)

102	review.ti. (867209)

103	99 and 102 (370138)

104	(review$ adj10 (papers or trials or trial data or studies or evidence or intervention$ or evaluation$ or outcome$ or findings)).ti,ab. (824317)

105	(retriev$ adj10 (papers or trials or studies or evidence or intervention$ or evaluation$ or outcome$ or findings)).ti,ab. (46542)

106	61 or 101 or 103 or 104 or 105 (1599909)

107	letter.pt. (1327839)

108	editorial.pt. (810830)

109	107 or 108 (2138669)

110	106 not 109 (1568161)

111	randomized controlled trial/ (828898)

112	controlled clinical trial/ (473459)

113	Random$.ti,ab,ot. (2085562)

114	randomization/ (99489)

115	intermethod comparison/ (307279)

116	placebo.ti,ab,ot. (379291)

117	(compare or compared or comparison).ti,ot. (628450)

118	((evaluated or evaluate or evaluating or assessed or assess) and (compare or compared or comparing or comparison)).ab. (2948399)

119	(open adj label).ti,ab,ot. (116367)

120	((double or single or doubly or singly) adj (blind or blinded or blindly)).ti,ab,ot. (283916)

121	double blind procedure/ (220584)

122	parallel group$1.ti,ab,ot. (33780)

123	(crossover or cross over).ti,ab,ot. (128969)

124	((assign$ or match or matched or allocation) adj5 (alternate or group or groups or intervention or interventions or patient or patients or subject or subjects or participant or participants)).ti,ab,ot. (436164)

125	(assigned or allocated).ti,ab,ot. (515550)

126	(controlled adj7 (study or design or trial)).ti,ab,ot. (475422)

127	(volunteer or volunteers).ti,ab,ot. (290432)

128	trial.ti,ot. (428134)

129	or/111-128 (6183681)

130	110 or 129 (7290857)

131	17 (409988)

132	130 and 131 (120720)

133	132 (120720)

134	(animal/ or animal experiment/ or animal model/ or animal tissue/ or nonhuman/) not exp human/ (7017940)

135	133 not 134 (119833)

136	133 and 134 (887)

137	(animal/ or nonhuman/) not exp human/ (6676639)

138	133 not 137 (119859)

139	133 and 137 (861)

140	animal/ or exp animal experiment/ or nonhuman/ (9784451)

141	(rat or rats or mouse or mice or hamster or hamsters or animal or animals or dog or dogs or cat or cats or bovine or sheep).ti,sh. (5707457)

142	or/140-141 (10345333)

143	exp human/ or human experiment/ (26722690)

144	142 not (142 and 143) (7385781)

145	133 not 144 (119789)

146	133 and 144 (931)

147	animal/ or exp animal experiment/ or nonhuman/ (9784451)

148	(rat or rats or mouse or mice or hamster or hamsters or animal or animals or dog or dogs or cat or cats or bovine or sheep).ti,ab,sh. (6681690)

149	or/147-148 (10749553)

150	exp human/ or human experiment/ (26722690)

151	149 not (149 and 150) (7521649)

152	133 not 151 (119746)

153	133 and 151 (974)

154	((animal or nonhuman) not human).sh. (6678852)

155	133 not 154 (119858)

156	133 and 154 (862)

157	animal experiment/ not (human experiment/ or human/) (2644485)

158	133 not 157 (120048)

159	133 and 157 (672)

160	(exp animal/ or exp invertebrate/ or nonhuman/ or animal experiment/ or animal tissue/ or animal model/ or exp plant/ or exp fungus/) not (exp human/ or human tissue/) (7956295)

161	133 not 160 (119701)

162	133 and 160 (1019)

163	(exp animal/ or exp invertebrate/ or nonhuman/ or animal experiment/ or animal tissue/ or animal model/ or exp plant/ or exp fungus/) not (exp human/ or human tissue/ or human experiment/) (7955132)

164	133 not 163 (119701)

165	133 and 163 (1019)

166	(exp animal/ or exp invertebrate/ or nonhuman/ or animal experiment/ or animal tissue/ or animal model/) not (exp human/ or human tissue/ or human experiment/) (7467817)

167	133 not 166 (119744)

168	133 and 166 (976)

169	(exp animals/ or exp animal experimentation/ or exp animal experiment/ or exp models animal/ or nonhuman/ or exp vertebrate/ or exp vertebrates/) not (exp humans/ or exp human experimentation/ or exp human experiment/) (7469930)

170	133 not 169 (119741)

171	133 and 169 (979)

172	(rat or rats or mouse or mice or swine or porcine or murine or sheep or lambs or pigs or piglets or rabbit or rabbits or cat or cats or dog or dogs or cattle or bovine or monkey or monkeys or trout or marmoset$1).ti. and animal experiment/ (1257365)

173	animal experiment/ not (human experiment/ or human/) (2644485)

174	or/172-173 (2719082)

175	133 not 174 (120013)

176	133 and 174 (707)

177	136 or 139 or 146 or 153 or 156 or 159 or 162 or 165 or 168 or 171 or 176 (1080)

178	135 or 138 or 145 or 152 or 155 or 158 or 161 or 164 or 167 or 170 or 175 (120057)

**Should be Removed (Animals)**

Embase via Ovid http://ovidsp.ovid.com/

Database Segment and Indexing: <1974 to 2024 November 19>

Date Searched: 20^th^ November 2024

1	("2010099353" or "638008172" or "640130493" or "2007492400" or "352761641" or "640479234" or "2022769579" or "619997060" or "2031291056" or "2011037848" or "2001707398" or "622599373" or "635482228" or "2024997694" or "2021252241" or "2020740833" or "644459240" or "2002120303" or "634191928" or "619032626" or "2007422405" or "2023770018" or "2007484477" or "2021230923" or "2004852123" or "2021765794" or "2004538792" or "2002247993" or "2032028099" or "2015702181" or "2003250919" or "625353258" or "2026489871" or "2010154625" or "643049053" or "2019435100" or "628507452" or "2000545774" or "2019463536" or "642120394" or "2028722341" or "2012092203" or "2019356796" or "2010203824" or "637516406" or "2001227748" or "634190907" or "2022468458" or "624431587" or "629412230" or "2020860531" or "2013013051" or "2013409346" or "2015571800" or "2020676059" or "629162486" or "2005885838" or "51654178" or "2005884056" or "2015314831" or "622722583" or "627534792" or "2022352994" or "2025302457" or "619681509" or "2018304670" or "628695937" or "2013841960" or "2032614636" or "2001850344" or "2014141191" or "635323558" or "635946301" or "2007333641" or "628867668" or "619483180" or "2014182318" or "629410767" or "629412364" or "642975066" or "628695963" or "629239234" or "2015785973" or "631198930" or "635426246" or "2011272352" or "2010550009" or "2011026247" or "627013692" or "2020260541" or "622352541" or "2002758418" or "627017173" or "2005163692" or "2026290034" or "2028093268" or "2030536394" or "2018816196" or "623038748" or "2018708319" or "631554698" or "2022317623" or "2023242921" or "624297866" or "2013514230" or "639870485" or "633592624" or "2001445755" or "622003069" or "364190764" or "633610806" or "640376565" or "2016449222" or "629411873" or "2010701709" or "629411408" or "629097615" or "635344322" or "636100875" or "637516656" or "629801846" or "2030304751" or "2028279168" or "2017252080" or "622637956" or "2028214902" or "2003987423" or "2029869326" or "629411883" or "2019780187" or "2007799211" or "2030463871" or "2015522697" or "2027256900" or "2000979915" or "2004128099" or "634314795" or "2015411001" or "637601233" or "2031572284" or "360231248" or "2010927725" or "2005426020" or "2023243126" or "2024614388" or "2013949274" or "2022818449" or "632996157" or "2011742743" or "2011823533" or "2021349852" or "2028430142" or "2028430142" or "627699151" or "633825224" or "635436082" or "618492322" or "2029703007" or "2000550563" or "2026523797" or "2021029795" or "629098204" or "640484134" or "2007241945" or "2026215738" or "2019877013" or "2030136123" or "2022663004" or "2026219920" or "629411958" or "634192736" or "634537689" or "632494146" or "2001349566" or "2028151712" or "2028858991" or "2024614287" or "2031556795" or "2002453746" or "629410813" or "629410924" or "624563971" or "358442034" or "627018462" or "2006921093" or "2011438374" or "628695988" or "2030841434" or "2022866076" or "2018084567" or "638356879" or "2020807152" or "2026491841" or "637752257" or "2029901989" or "2019769050" or "628902832" or "2015293919" or "631641571" or "2016445129" or "2001108648" or "2029642241" or "2028666617" or "2016470505" or "629410744" or "2015594234" or "2004258136" or "364943603" or "2001420355" or "2026213392" or "2017183089" or "22216183" or "624563894" or "622193014" or "631627090" or "2024837863" or "638323733" or "2018793555" or "2006994956" or "2004582739" or "626917143" or "639789843" or "2013098084" or "2016151556" or "2009011350" or "2016152123" or "2016151662" or "2008923566" or "2028767129" or "628240202" or "624843054" or "639227939" or "2003683311" or "2022307574" or "2018357976" or "2005621960" or "2027741277" or "2013913833" or "2006124295" or "643167812" or "2013731971" or "623004611" or "2020969831" or "644459125" or "2014612861" or "628309997" or "2003551533" or "2016027252" or "2005227916" or "352093768" or "2032712452" or "2029238944" or "2026596654" or "634191911" or "2001086689" or "625980507" or "632936877" or "641075904" or "637516377" or "623339287" or "639227909" or "2013840903" or "2028849949" or "2021980215" or "2003239343" or "635482280" or "2014925559" or "639526064" or "2024711906" or "638323777" or "2001879022" or "643356094" or "2013798958" or "2032433630" or "2024531166" or "638008510" or "2004483632" or "644344326" or "621915396" or "2029104457" or "2007748434" or "2030531682" or "2026575619" or "2000849860" or "2022133661" or "634149172" or "628554136" or "627211248" or "625510749" or "2004639909" or "2016800448" or "630726090" or "629416408" or "2021100812" or "2005487364" or "2004169818" or "2005548070" or "635753516" or "636274183" or "2000736540" or "638973599" or "629421942" or "624602560" or "2018075872" or "627649372" or "2016993839" or "2028389954" or "637515825" or "619934302" or "2016164587" or "2014698550" or "2019051384" or "2000947929").an. (312)

2	remove duplicates from 1 (312)

3	(animal/ or animal experiment/ or animal model/ or animal tissue/ or nonhuman/) not exp human/ (7127952)

4	2 not 3 (31)

5	2 and 3 (281)

6	(animal/ or nonhuman/) not exp human/ (6786390)

7	2 not 6 (31)

8	2 and 6 (281)

9	animal/ or exp animal experiment/ or nonhuman/ (9967558)

10	(rat or rats or mouse or mice or hamster or hamsters or animal or animals or dog or dogs or cat or cats or bovine or sheep).ti,sh. (5792466)

11	or/9-10 (10530178)

12	exp human/ or human experiment/ (27323434)

13	11 not (11 and 12) (7496062)

14	2 not 13 (30)

15	2 and 13 (282)

16	animal/ or exp animal experiment/ or nonhuman/ (9967558)

17	(rat or rats or mouse or mice or hamster or hamsters or animal or animals or dog or dogs or cat or cats or bovine or sheep).ti,ab,sh. (6782932)

18	or/16-17 (10939926)

19	exp human/ or human experiment/ (27323434)

20	18 not (18 and 19) (7633032)

21	2 not 20 (26)

22	2 and 20 (286)

23	((animal or nonhuman) not human).sh. (6788704)

24	2 not 23 (31)

25	2 and 23 (281)

26	animal experiment/ not (human experiment/ or human/) (2685603)

27	2 not 26 (83)

28	2 and 26 (229)

29	(exp animal/ or exp invertebrate/ or nonhuman/ or animal experiment/ or animal tissue/ or animal model/ or exp plant/ or exp fungus/) not (exp human/ or human tissue/) (8076067)

30	2 not 29 (27)

31	2 and 29 (285)

32	(exp animal/ or exp invertebrate/ or nonhuman/ or animal experiment/ or animal tissue/ or animal model/ or exp plant/ or exp fungus/) not (exp human/ or human tissue/ or human experiment/) (8074833)

33	2 not 32 (27)

34	2 and 32 (285)

35	(exp animal/ or exp invertebrate/ or nonhuman/ or animal experiment/ or animal tissue/ or animal model/) not (exp human/ or human tissue/ or human experiment/) (7578504)

36	2 not 35 (31)

37	2 and 35 (281)

38	(exp animals/ or exp animal experimentation/ or exp animal experiment/ or exp models animal/ or nonhuman/ or exp vertebrate/ or exp vertebrates/) not (exp humans/ or exp human experimentation/ or exp human experiment/) (7580901)

39	2 not 38 (29)

40	2 and 38 (283)

41	(rat or rats or mouse or mice or swine or porcine or murine or sheep or lambs or pigs or piglets or rabbit or rabbits or cat or cats or dog or dogs or cattle or bovine or monkey or monkeys or trout or marmoset$1).ti. and animal experiment/ (1275917)

42	animal experiment/ not (human experiment/ or human/) (2685603)

43	or/41-42 (2763141)

44	2 not 43 (61)

45	2 and 43 (251)

46	5 (281)

47	8 (281)

48	15 (282)

49	22 (286)

50	25 (281)

51	28 (229)

52	31 (285)

53	34 (285)

54	37 (281)

55	40 (283)

56	45 (251)

**Should be Retained (Not Animals)**

Embase via Ovid http://ovidsp.ovid.com/

Database Segment and Indexing: <1974 to 2024 November 19>

Date Searched: 20^th^ November 2024

1	("2004572601" or "2001405459" or "2001808382" or "2017145036" or "27261765" or "2021629005" or "624563910" or "643676676" or "2020587236" or "2032861656" or "624564732" or "2032867622" or "2022075667" or "644578408" or "2019150445" or "2020191906" or "2032982601" or "2030342196" or "2022548175" or "629302343" or "2020191306" or "2023737818" or "636981787" or "626434580" or "2007415928" or "2018693964" or "2016843031" or "2032855027" or "2019149777" or "2023737235" or "2032923850" or "2022548313" or "2020191590" or "643676777" or "2029562426" or "631118405" or "624431246" or "2030226531" or "2021043886" or "71947033" or "2017611595" or "2030784294" or "2017611594" or "2030338559" or "2032068856" or "2030243934" or "2016843021" or "2029529086" or "2017166815" or "644587290" or "2021629409" or "643676409" or "2029562135" or "623187887" or "2015874017" or "644562503" or "2001855108" or "2015722460" or "2027950551" or "2030322350" or "2032068313" or "2030274502" or "2028195100" or "70253318" or "2029517622" or "2019603011" or "2029895436" or "2017611156" or "2029518274" or "625158468" or "2021808000" or "2017166619" or "2018155143" or "2020586973" or "2022075518" or "2018694467" or "2016843452" or "626452684" or "2029775041" or "629411398" or "630319434" or "2021043299" or "2029466981" or "2023737335" or "2015873751" or "625343923" or "2016842966" or "2028841341" or "626758512" or "2013293812" or "624431756" or "2031182547" or "2031327700" or "2021629296" or "628695725" or "2017166713" or "2030314912" or "2032047497" or "2021629526" or "2016843570" or "2025489243" or "2020191876" or "2018154825" or "2024359322" or "2007804685" or "2021628969" or "2031673801" or "2026215134" or "2030219675" or "630320419" or "2020675525" or "2026215408" or "2017611365" or "2030219687" or "2026215133" or "2026945966" or "2004836513" or "2029140317" or "2024358715" or "2030025694" or "2027602611" or "2029869036" or "608115609" or "2028195018" or "2020191383" or "2030243854" or "2017677574" or "2011864415" or "2018423270" or "7113283" or "2017611020" or "628695406" or "2026214975" or "2017678969" or "2014259992" or "2029874938" or "2022548080" or "2017166776" or "2026945678" or "2030654945" or "2029517887" or "2027603516" or "627191017" or "2030219558" or "643676774" or "2017166249" or "2016370739" or "2030219674" or "2028427482" or "2023737562" or "2001203560" or "2024359373" or "2032069053" or "2011263619" or "644593555" or "2018693988" or "2025488559" or "2015874278" or "2018154590" or "2024974649" or "2030784110" or "2029840180" or "643574264" or "2022075591" or "29128277" or "2026945975" or "2028194233" or "2028778615" or "2031605050" or "2030266713" or "2026946041" or "644586386" or "2032819796" or "639393771" or "644564357" or "2004611240" or "2001493084" or "627135574" or "2018694563" or "2016370885" or "2030334400" or "2028194225" or "2026215337" or "623154921" or "2005627209" or "2012993679" or "2018154747" or "2001445858" or "2032068230" or "2016370415" or "2029517941" or "2029517941" or "2029775006" or "2024975256" or "2005043535" or "2020192017" or "2030326502" or "2030672513" or "2023147235" or "2031393982" or "2026945776" or "2030246907" or "2017166650" or "2028194858" or "2031135166" or "2024218404" or "644561470" or "2032867352" or "2032884331" or "624431310" or "624431615" or "2030348166" or "624431280" or "2025489013" or "644578814" or "2029809136" or "2019770681" or "2010599491" or "2010599488" or "2014389281" or "2016843684" or "630050130" or "2018154864" or "2023615273" or "2014023305" or "46106352" or "2017166413" or "2019602877" or "2024359021" or "2017611884" or "2032068138" or "2020587768" or "2015873743" or "2028194510" or "2015873897" or "644563349" or "2028778606" or "51889046" or "2018155067" or "2015873929" or "2029821280" or "2030285074" or "2032958009" or "2003529175" or "2031817883" or "2017166390" or "2013542514" or "2032937888" or "2028779274" or "2025149147" or "2017611846" or "2032955412" or "2028779352" or "2028778797" or "2029877847" or "2027602791" or "2032839169" or "2020223710" or "629309720" or "2030340002" or "2019150453" or "2032986354" or "2017166262" or "624714843" or "2023146808" or "2032902124" or "2019770506" or "629494810" or "2032838781" or "2032469465" or "611554644" or "2007088885" or "2030322701" or "2001511851" or "2029517709" or "2018694471" or "2029891050" or "2005178890" or "2018839210" or "2032866774" or "2030784151" or "627605583" or "2026214994" or "2022075707" or "350156070" or "2029518419" or "623173687" or "629427814" or "2023737319" or "626437797" or "2016370405" or "2022242632" or "2007239241" or "2032800038" or "2030303340" or "2024975157" or "2028014487" or "2027603402" or "2031596815" or "2003436349" or "2013184318" or "2023536854" or "2019150523" or "644576312" or "2030257601" or "2032838778" or "626460770" or "2031393997" or "2030285023" or "2021629433" or "2030173479" or "629412034" or "2030348675" or "2019150169" or "2002078048" or "644556730" or "2032932470" or "2029517713" or "2029877697" or "2031634533" or "2025488494" or "2014898143" or "2032910911" or "2030248317" or "2002912198" or "2022075622" or "2022548382" or "2030304063" or "626477948" or "2031394652" or "2030274550" or "2024395294" or "2032068721" or "626435471" or "2015511535" or "2029091360" or "2019150489" or "644588971" or "2030262495" or "2023737204" or "643676170" or "2030250761" or "2021043326" or "2028194717" or "2032914907" or "2026946169" or "2032774897" or "643364125" or "644564495" or "625863316" or "644584476" or "637040599" or "2030220067" or "2029655544" or "2019936653" or "2016843405" or "2020191579" or "2020587003" or "2023737483" or "2022548540" or "2020587370" or "2021629038" or "626516921" or "644587447" or "2019603154" or "2032941225" or "2029633608" or "2032844530" or "2017679583" or "2016199852" or "2030326893" or "624563897" or "2026892686" or "2030342063" or "2030284637" or "7088453" or "6191273" or "2030335951" or "2030897464" or "2029518043" or "624753299" or "626912187" or "2031596371" or "2030783458" or "2030342098" or "2032915013" or "2030250452" or "2016370511" or "2017581891" or "2023737693" or "2030307050" or "2027778123" or "2022075252" or "640429090" or "2018694304" or "2032884476" or "627251485" or "2025489109" or "2028194144" or "2017166051" or "2030199399" or "2006132160" or "9240656" or "2032942967" or "2001421801" or "620982075" or "623322732" or "2001401036" or "2016899791" or "2018019540" or "2032068217" or "636275965" or "2025503404" or "2027798801" or "2030896935" or "2026945774" or "2030322589" or "354655095" or "2021628919" or "2030270895" or "2027603359" or "2015874336" or "2021629300" or "627464487" or "2029623143" or "2014157754" or "2029517998" or "2032299368" or "2032942919" or "644556004" or "627190366" or "625572742" or "2030274988" or "2030783894" or "2032939635" or "638682177" or "2027603246" or "628710879" or "2024748212" or "2031594003" or "2032488833" or "2019150406" or "2030219820" or "2030783574" or "2008043484" or "2024814998" or "2014112693" or "2028194508" or "2030784054" or "2024359064" or "2027603423" or "623298294" or "2017166594" or "613272064" or "2031394010" or "2024975108" or "2031394940" or "636312895" or "2028194498" or "2023606587" or "29467080" or "600631152" or "636170043" or "2013951939" or "2030030570" or "2019602655" or "2032433852" or "626052682" or "2017613916" or "2017611189" or "2026946050" or "2016370733" or "644586446" or "2015039721" or "644576385" or "2017610866" or "624564924" or "2030143841" or "2020587042" or "2024975058" or "2026946475" or "2025488931" or "2018154786" or "2032821989" or "2025489014" or "2021629577" or "2030219275" or "2030231115" or "2015874126" or "2028214291" or "2010547152" or "2001445386" or "2019150167" or "2030219873" or "2015038998" or "2015873827" or "644563580" or "2023147229" or "2025488794" or "2030783433" or "2022075558" or "2030246424" or "2016370427" or "2026215414" or "2030187219" or "635342521" or "2030783419" or "624431117" or "626478658" or "2029935100" or "2015039229" or "2015511617" or "2030219575" or "2020191637" or "2031446121" or "2016843444" or "2024974930" or "2030346026" or "2020676082" or "2010153094" or "2032068921" or "2024975180" or "2030783620" or "2029455211" or "2029518029" or "2026945643" or "2020587193" or "628695947" or "2016080889" or "624431383" or "2026319565" or "2028194406" or "2032380475" or "2030326659" or "2021629501" or "2026945707" or "2019150395" or "70088192" or "2031394019" or "2023737603" or "2027952223" or "627463941" or "2024359117" or "2032821807" or "2030303390" or "2030783490" or "2031394150" or "2029273157" or "2029518335" or "2032859001" or "644555931" or "2030783474" or "2029930833" or "2017611851" or "2016897994" or "2031394509" or "2029905647" or "624732960" or "2016958049" or "2012508457" or "2029042375" or "2006923284" or "2017763483" or "2030219868" or "46902063" or "2031394403" or "2032069048" or "2017166852" or "2032859214" or "626456781" or "2023738813" or "50745671" or "618432880" or "630485871" or "2013583403" or "629783491" or "2015874207" or "2025001738" or "2024974636" or "2032879228" or "2001155219" or "2018694324" or "2029517783" or "2029748237" or "2002179641" or "2018155161" or "2031394512" or "2016370876" or "2019769273" or "2024974806" or "2030895419" or "2030243735" or "2024291614" or "642033997" or "2024397358" or "2029703044" or "2022075089" or "2016842997" or "2032346087" or "2032886317" or "2032939245" or "2032996730" or "2032891019" or "2018694548" or "2017166284" or "2023146671" or "2027187311" or "2029517994" or "2018694284" or "2027603001" or "2018694449" or "2016843667" or "2027603136" or "2018694217" or "2028779222" or "2030336357" or "2025488747" or "2032939674" or "2030784274" or "2007035540" or "623187625" or "359262404" or "624563880" or "2028194261" or "2030284645" or "2019150107" or "2022548526" or "2032914131" or "2030314957" or "2032987836" or "2017611212" or "2020191438" or "2024975241" or "2016843749" or "2030163948" or "2028194523" or "2028194891" or "2018154693" or "2030784229" or "602587040" or "2026945543" or "2007638320" or "2021394476" or "2032369817" or "2015873913" or "2023147078" or "623154909" or "2024359119" or "625667182" or "2027603030" or "2018155381" or "2030284565" or "2032866540" or "2027602581" or "2016816256" or "2031394896" or "2018760412" or "629410794" or "2032875800" or "2025675471" or "2026946096" or "2018839336" or "2030338787" or "2018154685" or "2015874018" or "2024974702" or "2024974702" or "2001401788" or "625666899" or "2007173764" or "630349504" or "2030783992" or "2017166143" or "2017611704" or "628240273" or "2030783912" or "2018215835" or "2022115046" or "2013616422" or "2022547822" or "620308849" or "2002953028" or "2030207206" or "2022075222" or "2001067459" or "643676516" or "2030831756").an. (678)

2	remove duplicates from 1 (678)

3	(animal/ or animal experiment/ or animal model/ or animal tissue/ or nonhuman/) not exp human/ (7127952)

4	2 not 3 (531)

5	2 and 3 (147)

6	(animal/ or nonhuman/) not exp human/ (6786390)

7	2 not 6 (532)

8	2 and 6 (146)

9	animal/ or exp animal experiment/ or nonhuman/ (9967558)

10	(rat or rats or mouse or mice or hamster or hamsters or animal or animals or dog or dogs or cat or cats or bovine or sheep).ti,sh. (5792466)

11	or/9-10 (10530178)

12	exp human/ or human experiment/ (27323434)

13	11 not (11 and 12) (7496062)

14	2 not 13 (530)

15	2 and 13 (148)

16	animal/ or exp animal experiment/ or nonhuman/ (9967558)

17	(rat or rats or mouse or mice or hamster or hamsters or animal or animals or dog or dogs or cat or cats or bovine or sheep).ti,ab,sh. (6782932)

18	or/16-17 (10939926)

19	exp human/ or human experiment/ (27323434)

20	18 not (18 and 19) (7633032)

21	2 not 20 (511)

22	2 and 20 (167)

23	((animal or nonhuman) not human).sh. (6788704)

24	2 not 23 (531)

25	2 and 23 (147)

26	animal experiment/ not (human experiment/ or human/) (2685603)

27	2 not 26 (599)

28	2 and 26 (79)

29	(exp animal/ or exp invertebrate/ or nonhuman/ or animal experiment/ or animal tissue/ or animal model/ or exp plant/ or exp fungus/) not (exp human/ or human tissue/) (8076067)

30	2 not 29 (520)

31	2 and 29 (158)

32	(exp animal/ or exp invertebrate/ or nonhuman/ or animal experiment/ or animal tissue/ or animal model/ or exp plant/ or exp fungus/) not (exp human/ or human tissue/ or human experiment/) (8074833)

33	2 not 32 (520)

34	2 and 32 (158)

35	(exp animal/ or exp invertebrate/ or nonhuman/ or animal experiment/ or animal tissue/ or animal model/) not (exp human/ or human tissue/ or human experiment/) (7578504)

36	2 not 35 (530)

37	2 and 35 (148)

38	(exp animals/ or exp animal experimentation/ or exp animal experiment/ or exp models animal/ or nonhuman/ or exp vertebrate/ or exp vertebrates/) not (exp humans/ or exp human experimentation/ or exp human experiment/) (7580901)

39	2 not 38 (529)

40	2 and 38 (149)

41	(rat or rats or mouse or mice or swine or porcine or murine or sheep or lambs or pigs or piglets or rabbit or rabbits or cat or cats or dog or dogs or cattle or bovine or monkey or monkeys or trout or marmoset$1).ti. and animal experiment/ (1275917)

42	animal experiment/ not (human experiment/ or human/) (2685603)

43	or/41-42 (2763141)

44	2 not 43 (589)

45	2 and 43 (89)

46	4 (531)

47	7 (532)

48	14 (530)

49	21 (511)

50	24 (531)

51	27 (599)

52	30 (520)

53	33 (520)

54	36 (530)

55	39 (529)

56	44 (589)

### **Dataset – Animals/Nonhumans and Humans:**

#### **Lines to Retrieve Records for Animals/Nonhumans and Humans Dataset**

Embase via Ovid http://ovidsp.ovid.com/

Database Segment and Indexing: <1974 to 2024 June 28>

Date Searched: 27^th^ June 2024

1	anthrozoolog*.af. (280)

2	human-animal bond*.af. (1092)

3	animal-human bond*.af. (14)

4	cat scratch disease.af. (3328)

5	bartonella henselae.af. (3204)

6	xenograft*.af. (167450)

7	"guide dog".af. (132)

8	police horse.af. (4)

9	police dog.af. (108)

10	cadaver dogs.af. (67)

11	animal-assisted therapy.af. (1044)

12	zoonotic disease*.af. (12273)

13	zoonoses.af. (19422)

14	human-animal chimera*.af. (63)

15	(giant hogweed adj4 burn*).af. (7)

16	aspergillosis.af. (35499)

17	or/1-16 (240240)

18	systematic$ review$.ti,ab. (410334)

19	systematic$ literature review$.ti,ab. (28631)

20	"systematic review"/ (473545)

21	"systematic review (topic)"/ (35217)

22	meta analysis/ (320342)

23	"meta analysis (topic)"/ (56278)

24	meta-analytic$.ti,ab. (12292)

25	meta-analysis.ti,ab. (333845)

26	metanalysis.ti,ab. (1485)

27	metaanalysis.ti,ab. (10710)

28	meta analysis.ti,ab. (333845)

29	meta-synthesis.ti,ab. (1904)

30	metasynthesis.ti,ab. (569)

31	meta synthesis.ti,ab. (1904)

32	meta-regression.ti,ab. (17851)

33	metaregression.ti,ab. (1589)

34	meta regression.ti,ab. (17851)

35	(synthes$ adj3 literature).ti,ab. (7512)

36	(synthes$ adj3 evidence).ti,ab. (22111)

37	(synthes$ adj2 qualitative).ti,ab. (7555)

38	integrative review.ti,ab. (5422)

39	data synthesis.ti,ab. (17753)

40	(research synthesis or narrative synthesis).ti,ab. (9265)

41	(systematic study or systematic studies).ti,ab. (17095)

42	(systematic comparison$ or systematic overview$).ti,ab. (5552)

43	(systematic adj2 search$).ti,ab. (62464)

44	systematic$ literature research$.ti,ab. (543)

45	(review adj3 scientific literature).ti,ab. (2897)

46	(literature review adj2 side effect$).ti,ab. (24)

47	(literature review adj2 adverse effect$).ti,ab. (10)

48	(literature review adj2 adverse event$).ti,ab. (21)

49	(evidence-based adj2 review).ti,ab. (4969)

50	comprehensive review.ti,ab. (33841)

51	critical review.ti,ab. (21992)

52	critical analysis.ti,ab. (10761)

53	quantitative review.ti,ab. (1007)

54	structured review.ti,ab. (1455)

55	realist review.ti,ab. (757)

56	realist synthesis.ti,ab. (402)

57	(pooled adj2 analysis).ti,ab. (33479)

58	(pooled data adj6 (studies or trials)).ti,ab. (4316)

59	(medline and (inclusion adj3 criteria)).ti,ab. (41638)

60	(search adj (strateg$ or term$)).ti,ab. (60127)

61	or/18-60 (945654)

62	medline.ab. (222986)

63	pubmed.ab. (296450)

64	cochrane.ab. (190840)

65	embase.ab. (217402)

66	cinahl.ab. (58572)

67	psyc?lit.ab. (1012)

68	psyc?info.ab. (62148)

69	lilacs.ab. (12574)

70	(literature adj3 search$).ab. (128249)

71	(database$ adj3 search$).ab. (134000)

72	(bibliographic adj3 search$).ab. (4961)

73	(electronic adj3 search$).ab. (44060)

74	(electronic adj3 database$).ab. (68208)

75	(computeri?ed adj3 search$).ab. (5026)

76	(internet adj3 search$).ab. (6095)

77	included studies.ab. (62285)

78	(inclusion adj3 studies).ab. (34855)

79	inclusion criteria.ab. (243967)

80	selection criteria.ab. (47347)

81	predefined criteria.ab. (3712)

82	predetermined criteria.ab. (1718)

83	(assess$ adj3 (quality or validity)).ab. (163525)

84	(select$ adj3 (study or studies)).ab. (128134)

85	(data adj3 extract$).ab. (148505)

86	extracted data.ab. (30123)

87	(data adj2 abstracted).ab. (12241)

88	(data adj3 abstraction).ab. (3736)

89	published intervention$.ab. (331)

90	((study or studies) adj2 evaluat$).ab. (383830)

91	(intervention$ adj2 evaluat$).ab. (23090)

92	confidence interval$.ab. (739122)

93	heterogeneity.ab. (331257)

94	pooled.ab. (195113)

95	pooling.ab. (23675)

96	odds ratio$.ab. (500130)

97	(Jadad or coding).ab. (289769)

98	evidence-based.ti,ab. (213603)

99	or/62-98 (3028500)

100	review.pt. (3243148)

101	99 and 100 (421773)

102	review.ti. (867209)

103	99 and 102 (370138)

104	(review$ adj10 (papers or trials or trial data or studies or evidence or intervention$ or evaluation$ or outcome$ or findings)).ti,ab. (824317)

105	(retriev$ adj10 (papers or trials or studies or evidence or intervention$ or evaluation$ or outcome$ or findings)).ti,ab. (46542)

106	61 or 101 or 103 or 104 or 105 (1599909)

107	letter.pt. (1327839)

108	editorial.pt. (810830)

109	107 or 108 (2138669)

110	106 not 109 (1568161)

111	randomized controlled trial/ (828898)

112	controlled clinical trial/ (473459)

113	Random$.ti,ab,ot. (2085562)

114	randomization/ (99489)

115	intermethod comparison/ (307279)

116	placebo.ti,ab,ot. (379291)

117	(compare or compared or comparison).ti,ot. (628450)

118	((evaluated or evaluate or evaluating or assessed or assess) and (compare or compared or comparing or comparison)).ab. (2948399)

119	(open adj label).ti,ab,ot. (116367)

120	((double or single or doubly or singly) adj (blind or blinded or blindly)).ti,ab,ot. (283916)

121	double blind procedure/ (220584)

122	parallel group$1.ti,ab,ot. (33780)

123	(crossover or cross over).ti,ab,ot. (128969)

124	((assign$ or match or matched or allocation) adj5 (alternate or group or groups or intervention or interventions or patient or patients or subject or subjects or participant or participants)).ti,ab,ot. (436164)

125	(assigned or allocated).ti,ab,ot. (515550)

126	(controlled adj7 (study or design or trial)).ti,ab,ot. (475422)

127	(volunteer or volunteers).ti,ab,ot. (290432)

128	trial.ti,ot. (428134)

129	or/111-128 (6183681)

130	110 or 129 (7290857)

131	17 (240240)

132	130 and 131 (39898)

133	132 (39898)

134	(animal/ or animal experiment/ or animal model/ or animal tissue/ or nonhuman/) not exp human/ (7017940)

135	133 not 134 (30732)

136	133 and 134 (9166)

137	(animal/ or nonhuman/) not exp human/ (6676639)

138	133 not 137 (30832)

139	133 and 137 (9066)

140	animal/ or exp animal experiment/ or nonhuman/ (9784451)

141	(rat or rats or mouse or mice or hamster or hamsters or animal or animals or dog or dogs or cat or cats or bovine or sheep).ti,sh. (5707457)

142	or/140-141 (10345333)

143	exp human/ or human experiment/ (26722690)

144	142 not (142 and 143) (7385781)

145	133 not 144 (30062)

146	133 and 144 (9836)

147	animal/ or exp animal experiment/ or nonhuman/ (9784451)

148	(rat or rats or mouse or mice or hamster or hamsters or animal or animals or dog or dogs or cat or cats or bovine or sheep).ti,ab,sh. (6681690)

149	or/147-148 (10749553)

150	exp human/ or human experiment/ (26722690)

151	149 not (149 and 150) (7521649)

152	133 not 151 (29801)

153	133 and 151 (10097)

154	((animal or nonhuman) not human).sh. (6678852)

155	133 not 154 (30811)

156	133 and 154 (9087)

157	animal experiment/ not (human experiment/ or human/) (2644485)

158	133 not 157 (33261)

159	133 and 157 (6637)

160	(exp animal/ or exp invertebrate/ or nonhuman/ or animal experiment/ or animal tissue/ or animal model/ or exp plant/ or exp fungus/) not (exp human/ or human tissue/) (7956295)

161	133 not 160 (29805)

162	133 and 160 (10093)

163	(exp animal/ or exp invertebrate/ or nonhuman/ or animal experiment/ or animal tissue/ or animal model/ or exp plant/ or exp fungus/) not (exp human/ or human tissue/ or human experiment/) (7955132)

164	133 not 163 (29806)

165	133 and 163 (10092)

166	(exp animal/ or exp invertebrate/ or nonhuman/ or animal experiment/ or animal tissue/ or animal model/) not (exp human/ or human tissue/ or human experiment/) (7467817)

167	133 not 166 (29894)

168	133 and 166 (10004)

169	(exp animals/ or exp animal experimentation/ or exp animal experiment/ or exp models animal/ or nonhuman/ or exp vertebrate/ or exp vertebrates/) not (exp humans/ or exp human experimentation/ or exp human experiment/) (7469930)

170	133 not 169 (29876)

171	133 and 169 (10022)

172	(rat or rats or mouse or mice or swine or porcine or murine or sheep or lambs or pigs or piglets or rabbit or rabbits or cat or cats or dog or dogs or cattle or bovine or monkey or monkeys or trout or marmoset$1).ti. and animal experiment/ (1257365)

173	animal experiment/ not (human experiment/ or human/) (2644485)

174	or/172-173 (2719082)

175	133 not 174 (31862)

176	133 and 174 (8036)

177	136 or 139 or 146 or 153 or 156 or 159 or 162 or 165 or 168 or 171 or 176 (11654)

178	135 or 138 or 145 or 152 or 155 or 158 or 161 or 164 or 167 or 170 or 175 (33329)

**Should be Removed (Animals)**

Embase via Ovid http://ovidsp.ovid.com/

Database Segment and Indexing: <1974 to 2024 November 19>

Date Searched: 20^th^ November 2024

1	("2031588078" or "2029870952" or "2032612296" or "2031042234" or "2028039834" or "2032756968" or "2030859110" or "633152780" or "2029406836" or "644243352" or "644243943" or "643077197" or "643163440" or "2023573378" or "2029265447" or "2030036029" or "2026483677" or "2027114953" or "2030109984" or "2028613468" or "628981055" or "2032026810" or "644391546" or "607379208" or "633300706" or "72195767" or "2028103596" or "632261292" or "643076731" or "629323354" or "2026988934" or "644078479" or "644243430" or "643358101" or "2029270770" or "2011987947" or "2030006242" or "643177208" or "2028787901" or "644243971" or "2028418703" or "629337524" or "644029273" or "2010196854" or "621926276" or "2032759449" or "2032118231" or "2029406790" or "2030895396" or "2029868100" or "2022243574" or "71652350" or "2031748033" or "644223540" or "2027301313" or "600635460" or "2028523464" or "644242023" or "2024392769" or "629324606" or "644244276" or "2028726750" or "643823281" or "644242181" or "2029869327" or "2028608502" or "2028747040" or "2029685677" or "2028913116" or "2008404938" or "643160272" or "2031591835" or "644243984" or "2031346074" or "2031085318" or "2031547367" or "2018218130" or "2023327078" or "2027008605" or "644243981" or "630988811" or "2024396710" or "644242898" or "643161153" or "2032065879" or "2028626422" or "2028365202" or "2032369689" or "644243874" or "630548479" or "644243877" or "2025205941" or "2028870880" or "630180222" or "644313546" or "612964903" or "2029986565" or "625975639" or "2027335431" or "2032171604" or "644241618" or "2030046703" or "2031732506" or "2032153758" or "2031654099" or "644504945" or "644239657" or "2028907791" or "643076369" or "2028662206" or "2032706110" or "643050010" or "2030293817" or "644562074" or "2032596020" or "2029634304" or "644239846" or "644253309" or "2026319785" or "643569555" or "643076363" or "629323728" or "2028642921" or "643076245" or "2029135397" or "2029344338" or "2032548335" or "644240742" or "2030314334" or "629403342" or "2022241260" or "625347280" or "2032950443" or "2016899507" or "2023425652" or "643130446" or "2030800957" or "644313421" or "2032463473" or "2030130046" or "625864181" or "2029777044" or "2028765526" or "2031582445" or "2029621222" or "644237443" or "35001479" or "2030035355" or "2031823677" or "644314556" or "2023615521" or "2017266547" or "2028690967" or "2031281748" or "2015898602" or "2029466077" or "364186846" or "644242784" or "2027984207" or "2029055505" or "2024397845" or "644313637" or "644223132" or "2028872172" or "2028774183" or "644243964" or "644239231" or "644238637" or "2028608763" or "644239757" or "644314811" or "631296291" or "2024392989" or "2028194304" or "2025675256" or "644243466" or "2029472328" or "2024999829" or "2029803113" or "634141673" or "629438749" or "600544895" or "643977839" or "2032145333" or "2023326210" or "2028733881" or "2029420348" or "2018217848" or "644243600" or "644242548" or "2015568501" or "644244398" or "644243396" or "644228555" or "2018832262" or "2028884635" or "2030705446" or "2032343652" or "2023327209" or "2028526065" or "2027952371" or "2016900355" or "2031253551" or "2032433781" or "643076498" or "2016405010" or "644440701" or "2029665645" or "2028838304" or "2031192568" or "2029924367" or "2032294316" or "2030109634" or "644313650" or "29288368" or "2028215188" or "644313983" or "2029119141" or "2028803323" or "644243661" or "644314752" or "2027950608" or "644239394" or "644243689" or "644181321" or "2029712868" or "629010973" or "2030248799" or "643783390" or "644313956" or "2028430423" or "2032628482" or "2030109544" or "2029528416" or "644243109" or "2032678794" or "2027657900" or "2028871670" or "2015549628" or "2029407210" or "643158312" or "53159749" or "644506051" or "2029943963" or "51814170" or "644314194" or "2030198711" or "2029353525" or "2029448696" or "2020768589" or "2031929837" or "2027949916" or "625975206" or "2030908934" or "2032637515" or "2015896616" or "644223638" or "2029860956" or "2029794902" or "629324670" or "644242175" or "644222844" or "644457686" or "2031733383" or "2032027782" or "2024392111" or "2028685554" or "2027323731" or "644314377" or "644396264" or "2031905478" or "2017398938" or "637069188" or "2031330062" or "2030322549" or "2029839131" or "644242273" or "644314340" or "2014450048" or "2029215167" or "2031594385" or "29453498" or "2029984540" or "2017611067" or "644242642" or "605783997" or "643443175" or "644242506" or "2032013180" or "644241208" or "2028175802" or "2025673808" or "2032907310" or "2031668020" or "2032558939" or "644314804" or "2029823734" or "642518451" or "2032195225" or "2029991188" or "2030714130" or "644396491" or "1370245475" or "633847249" or "2031324866" or "2030863550" or "2028861012" or "643121838" or "643424784" or "2028599414" or "644243515" or "644243917" or "625971543" or "643299845" or "644440364" or "2028626779" or "2032579382" or "644339409" or "2029377464" or "644405752" or "2030395942" or "2022408685" or "2013840097" or "644298205" or "644243754" or "2028510558" or "2029550515" or "2027819177" or "644313919" or "637085209" or "2027008458" or "625969852" or "2030774316" or "2030173868" or "644244209" or "2029846490" or "643163312" or "611068821" or "2030348937" or "2030199117" or "2028715023" or "643468869" or "2028761369" or "644576808" or "2027917876" or "2028886137" or "2029875850" or "2028052755" or "2030814551" or "644313903" or "2030722727" or "629323527" or "2027777454" or "644242807" or "2030057397" or "644589229" or "644244170" or "614310454" or "2027198441" or "2029837062" or "2032094619" or "2032010838" or "643482752" or "630583122" or "644546714" or "644455799" or "2032891234" or "2029545940" or "2032871478" or "2032589793" or "2030136209" or "2029040533" or "2029875931" or "644243510" or "625346633" or "2031909553" or "2032050672" or "2015898642" or "644243181" or "2028744045" or "644243132" or "644242607" or "644458073" or "644241672" or "2026584004" or "2018217339" or "2029717926" or "644242144" or "2029938714" or "71760974" or "644242025" or "644243476" or "2027951167" or "644243376" or "644241058" or "643077065" or "1358460588" or "2030268918" or "635573009" or "644241601" or "2029209704" or "2032212101" or "2020092247" or "644243672" or "644242461" or "644243171" or "644242554" or "53163718" or "644243912" or "2027735115" or "2031294471" or "644243366" or "372646611" or "625971813" or "643088143" or "643160316" or "644239054" or "2028430163" or "644243833" or "2028523844" or "2029282388" or "2028732047" or "2026958391" or "644243010" or "2032011739" or "2029487292" or "1358354322" or "2032897393" or "644343967" or "642138012" or "2027205832" or "644056579" or "644052035" or "644266144" or "2028747782" or "2020674776" or "642825448" or "643051711" or "644242919" or "644242987" or "2031993401" or "644441323" or "638443999" or "2028783099" or "644243944" or "643321598" or "2032108921" or "2028106631" or "643121951" or "644240708" or "2030104425" or "2029588275" or "71671489" or "644242875" or "2031865267" or "2028837973" or "2028601108" or "2029841105" or "643049969" or "2015897838" or "644132140" or "2028884462" or "644242612" or "644259536" or "2030122320" or "2029153310" or "631071008" or "2029891286" or "644514579" or "2028316202" or "633993462" or "2030318994" or "643169312" or "2029800674" or "2028933703" or "2024835706" or "2021263364" or "2026888297" or "2028597096" or "2032299064" or "2029731632" or "2028939530" or "2032951723" or "643673244" or "2022781986" or "2032118252" or "643804700" or "629439611" or "644239488" or "644241600" or "71522930" or "2029806274" or "625968240" or "644339361" or "2017267636" or "2024880853" or "2017678112" or "2017678049" or "2019455806" or "2031910641" or "644241667" or "644314619" or "2032027773" or "2027070199" or "620424172" or "2030622267" or "644244264" or "2029567052" or "644243227" or "2029047188" or "2032649919" or "616994099" or "2032384875" or "642833900" or "644242728" or "643477733" or "643050051" or "2029984973" or "2032932190" or "2030916802" or "2031997517" or "643161126" or "2032154349" or "2029369018" or "2029820210" or "2026260760" or "643770448" or "2031042219" or "643160365" or "2028123329" or "2032681565" or "2028293575" or "2030138119" or "2029834554" or "2029718084" or "2032217543" or "625347991" or "2004374575" or "644238737" or "644243222" or "644095408" or "2030153611" or "2029282382" or "2029353523" or "2031923483" or "2028871708" or "2026766859" or "369034259" or "644242871" or "71762880" or "2032678523" or "644587508" or "2031836543" or "2025132860" or "2032596018" or "372175842" or "643461325" or "644243653" or "626832170" or "644223604" or "2031634103" or "2029991460" or "2025230159" or "629323862" or "2032434894" or "644240486" or "2028139612" or "2028743887" or "2028595824" or "2029986016" or "2032909555" or "643392959" or "2016437260" or "2032121476" or "629323555" or "644243129" or "2027866898" or "2024395038" or "2028774463" or "2032212892" or "644314961" or "644242193" or "2028655613" or "637041698" or "2031345473" or "2029824588" or "2028595508" or "644331552" or "607819043" or "2029148986" or "2027721950" or "644242297" or "616838781" or "2022243095" or "2030894442" or "2028750910" or "2029994508" or "643783763" or "2029878344" or "2032915170" or "2028520829" or "2031467619" or "2030395226" or "2030932929" or "2029985867" or "629323534" or "2024398144" or "2028876756" or "2025674497" or "629316464" or "2032128303" or "2028872202" or "2027984822" or "644378084" or "644240816" or "2028083063" or "2028743402" or "2027005359" or "644238579" or "2025786345" or "2029501980" or "2028525472" or "643444182" or "2028859198" or "2028859199" or "2032017717" or "2029577999" or "644242442" or "2029892189" or "638934001" or "640999440" or "644243295" or "2032018749" or "2032335658" or "644347694" or "2030039199" or "633826218" or "2004472987" or "2030269784" or "2032423902" or "2028933385" or "2017704613" or "644514346" or "644166870" or "644242786" or "2032027843" or "644444048" or "2029598967" or "644242429" or "634433179" or "625348093" or "2031853692" or "2032202324" or "2029949918" or "2022042262" or "2032068173" or "2030215758" or "2029693137" or "628125871" or "2028935826" or "2027970884" or "644243809").an. (652)

2	remove duplicates from 1 (652)

3	(animal/ or animal experiment/ or animal model/ or animal tissue/ or nonhuman/) not exp human/ (7127952)

4	2 not 3 (320)

5	2 and 3 (332)

6	(animal/ or nonhuman/) not exp human/ (6786390)

7	2 not 6 (320)

8	2 and 6 (332)

9	animal/ or exp animal experiment/ or nonhuman/ (9967558)

10	(rat or rats or mouse or mice or hamster or hamsters or animal or animals or dog or dogs or cat or cats or bovine or sheep).ti,sh. (5792466)

11	or/9-10 (10530178)

12	exp human/ or human experiment/ (27323434)

13	11 not (11 and 12) (7496062)

14	2 not 13 (318)

15	2 and 13 (334)

16	animal/ or exp animal experiment/ or nonhuman/ (9967558)

17	(rat or rats or mouse or mice or hamster or hamsters or animal or animals or dog or dogs or cat or cats or bovine or sheep).ti,ab,sh. (6782932)

18	or/16-17 (10939926)

19	exp human/ or human experiment/ (27323434)

20	18 not (18 and 19) (7633032)

21	2 not 20 (281)

22	2 and 20 (371)

23	((animal or nonhuman) not human).sh. (6788704)

24	2 not 23 (319)

25	2 and 23 (333)

26	animal experiment/ not (human experiment/ or human/) (2685603)

27	2 not 26 (413)

28	2 and 26 (239)

29	(exp animal/ or exp invertebrate/ or nonhuman/ or animal experiment/ or animal tissue/ or animal model/ or exp plant/ or exp fungus/) not (exp human/ or human tissue/) (8076067)

30	2 not 29 (322)

31	2 and 29 (330)

32	(exp animal/ or exp invertebrate/ or nonhuman/ or animal experiment/ or animal tissue/ or animal model/ or exp plant/ or exp fungus/) not (exp human/ or human tissue/ or human experiment/) (8074833)

33	2 not 32 (322)

34	2 and 32 (330)

35	(exp animal/ or exp invertebrate/ or nonhuman/ or animal experiment/ or animal tissue/ or animal model/) not (exp human/ or human tissue/ or human experiment/) (7578504)

36	2 not 35 (322)

37	2 and 35 (330)

38	(exp animals/ or exp animal experimentation/ or exp animal experiment/ or exp models animal/ or nonhuman/ or exp vertebrate/ or exp vertebrates/) not (exp humans/ or exp human experimentation/ or exp human experiment/) (7580901)

39	2 not 38 (321)

40	2 and 38 (331)

41	(rat or rats or mouse or mice or swine or porcine or murine or sheep or lambs or pigs or piglets or rabbit or rabbits or cat or cats or dog or dogs or cattle or bovine or monkey or monkeys or trout or marmoset$1).ti. and animal experiment/ (1275917)

42	animal experiment/ not (human experiment/ or human/) (2685603)

43	or/41-42 (2763141)

44	2 not 43 (370)

45	2 and 43 (282)

46	5 (332)

47	8 (332)

48	15 (334)

49	22 (371)

50	25 (333)

51	28 (239)

52	31 (330)

53	34 (330)

54	37 (330)

55	40 (331)

56	45 (282)

**Should be Retained (Not Animals)**

Embase via Ovid http://ovidsp.ovid.com/

Database Segment and Indexing: <1974 to 2024 November 19>

Date Searched: 20^th^ November 2024

1	("2016842768" or "625343769" or "2029291482" or "629325082" or "2031711326" or "610669609" or "2023328117" or "644284071" or "629337866" or "2024974702" or "604426625" or "625977407" or "615253305" or "625347283" or "2026180049" or "2013601213" or "644447488" or "2029646435" or "644382975" or "2028423884" or "2031148060" or "2027666977" or "2028838288" or "2029442073" or "643070680" or "2026191461" or "2030284835" or "2031334625" or "2030203039" or "2030236196" or "2029211376" or "2028658183" or "2027750124" or "643899444" or "2008490664" or "2026811171" or "2029621180" or "2029813254" or "629405131" or "2028872376" or "623683500" or "2029221714" or "2029815075" or "2031291554" or "2032771772" or "2029559690" or "2030571049" or "2018311827" or "2028011077" or "2028585449" or "2028037446" or "2029489478" or "2028979836" or "2029949483" or "2029225400" or "2028886256" or "2029214391" or "2004085360" or "632628017" or "2028825860" or "2028059086" or "2032111271" or "2031889102" or "644098429" or "2032735621" or "2032283215" or "2027348847" or "2028904923" or "2027948706" or "2030727506" or "2029161891" or "2029771314" or "2027959924" or "2023019688" or "624200522" or "2030153952" or "2025173222" or "2025000686" or "2030932779" or "2032038663" or "617374161" or "613548076" or "2030048131" or "373252075" or "2029703009" or "2029894601" or "2032718273" or "2026712696" or "2032347707" or "2030064565" or "2032248300" or "2032677946" or "2016438657" or "2032583027" or "2032685122" or "2032771169" or "2016842768" or "2029892367" or "2032613524" or "2032454907" or "644440354" or "2032776813" or "2032395002" or "644241685" or "2029646435" or "2030243993" or "2028974500" or "2029937784" or "644382975" or "2030038908" or "2032354278" or "2030020589" or "2032859227" or "2032789358" or "644265847" or "2030834936" or "644436004" or "2028460176" or "2032068772" or "2022547860" or "2031082477" or "2032492490" or "2026215084" or "644561863" or "2021629015" or "2029585738" or "2031971346" or "644382781" or "643260981" or "2022075345" or "2032150753" or "644243235" or "2030956632" or "644261081" or "2032681611" or "2023737672" or "644441798" or "2029877721" or "2030284835" or "2027156881" or "644242441" or "2017611133" or "644427631" or "2029353042" or "2029678228" or "2032429194" or "644584366" or "2025487593" or "644314905" or "2029863535" or "2032859207" or "2031711326" or "2030061894" or "2029646454" or "2032394965" or "644396776" or "2032452450" or "2029247586" or "2029867873" or "2029646427" or "2031394660" or "643906291" or "2031889102" or "644314834" or "2022075841" or "2032652581" or "2030236196" or "2030173443" or "644314802" or "2032501626" or "2032735621" or "2032867261" or "2032283215" or "2028194526" or "2030174267" or "2017611289" or "2032953633" or "2026320351" or "2032452604" or "2028491452" or "2029646471" or "643899444" or "2030181076" or "2025002246" or "2030235843" or "2032393866" or "627237353" or "644257607" or "2030074931" or "644440855" or "644586392" or "644439409" or "2032811133" or "2023328117" or "644589683" or "2029688480" or "2032896805" or "644283613" or "2030114419" or "2029771314" or "2008490664" or "2030073163" or "2032146773" or "2029870186" or "2029647007" or "2032492463" or "2032613525" or "2032418544" or "2030905072" or "2032069029" or "2032560950" or "644314070" or "2029732206" or "2027113897" or "2032894719" or "644284071" or "2030057439" or "2026889279" or "2032859214" or "2028778829" or "2032669829" or "2029295086" or "2030033367" or "2029813254" or "644314565" or "2032775239" or "2029730089" or "2029871403" or "2022693608" or "2026490434" or "2024974702" or "2024974751" or "644314162" or "2032047715" or "2029949483" or "2029603363" or "2032068906" or "643803020" or "2029066885" or "2030783704" or "2023019688" or "2030783932" or "2032623800" or "2029517964" or "644514480" or "2029815075" or "644522222" or "2030244223" or "644326045" or "2032392197" or "2031634594" or "2030221597" or "2032601537" or "644520682" or "2018694238" or "2029270720" or "2029742940" or "644391234" or "2028644292" or "2030303425" or "643616638" or "2020674826" or "644344183" or "644435842" or "2029874481" or "2025000686" or "2018155232" or "2030303156" or "2019150225" or "2030274563" or "644441232" or "644269645" or "2031594983" or "2032950631" or "2030783630" or "644524623" or "2016030860" or "2032200741" or "2016371240" or "644266176" or "2015548200" or "2028928211" or "644280937" or "2032211358" or "2026215051" or "644313859" or "2030262901" or "2032111170" or "642789249" or "2032273739" or "644242237" or "644314068" or "2019602712" or "2029397074" or "2032188827" or "2030104247" or "644242186" or "644334816" or "2032706468" or "644447488" or "2030285176" or "2015896454" or "2029413571" or "2032953590" or "2029481362").an. (273)

2	remove duplicates from 1 (273)

3	(animal/ or animal experiment/ or animal model/ or animal tissue/ or nonhuman/) not exp human/ (7127952)

4	2 not 3 (209)

5	2 and 3 (64)

6	(animal/ or nonhuman/) not exp human/ (6786390)

7	2 not 6 (210)

8	2 and 6 (63)

9	animal/ or exp animal experiment/ or nonhuman/ (9967558)

10	(rat or rats or mouse or mice or hamster or hamsters or animal or animals or dog or dogs or cat or cats or bovine or sheep).ti,sh. (5792466)

11	or/9-10 (10530178)

12	exp human/ or human experiment/ (27323434)

13	11 not (11 and 12) (7496062)

14	2 not 13 (209)

15	2 and 13 (64)

16	animal/ or exp animal experiment/ or nonhuman/ (9967558)

17	(rat or rats or mouse or mice or hamster or hamsters or animal or animals or dog or dogs or cat or cats or bovine or sheep).ti,ab,sh. (6782932)

18	or/16-17 (10939926)

19	exp human/ or human experiment/ (27323434)

20	18 not (18 and 19) (7633032)

21	2 not 20 (198)

22	2 and 20 (75)

23	((animal or nonhuman) not human).sh. (6788704)

24	2 not 23 (210)

25	2 and 23 (63)

26	animal experiment/ not (human experiment/ or human/) (2685603)

27	2 not 26 (256)

28	2 and 26 (17)

29	(exp animal/ or exp invertebrate/ or nonhuman/ or animal experiment/ or animal tissue/ or animal model/ or exp plant/ or exp fungus/) not (exp human/ or human tissue/) (8076067)

30	2 not 29 (208)

31	2 and 29 (65)

32	(exp animal/ or exp invertebrate/ or nonhuman/ or animal experiment/ or animal tissue/ or animal model/ or exp plant/ or exp fungus/) not (exp human/ or human tissue/ or human experiment/) (8074833)

33	2 not 32 (208)

34	2 and 32 (65)

35	(exp animal/ or exp invertebrate/ or nonhuman/ or animal experiment/ or animal tissue/ or animal model/) not (exp human/ or human tissue/ or human experiment/) (7578504)

36	2 not 35 (208)

37	2 and 35 (65)

38	(exp animals/ or exp animal experimentation/ or exp animal experiment/ or exp models animal/ or nonhuman/ or exp vertebrate/ or exp vertebrates/) not (exp humans/ or exp human experimentation/ or exp human experiment/) (7580901)

39	2 not 38 (207)

40	2 and 38 (66)

41	(rat or rats or mouse or mice or swine or porcine or murine or sheep or lambs or pigs or piglets or rabbit or rabbits or cat or cats or dog or dogs or cattle or bovine or monkey or monkeys or trout or marmoset$1).ti. and animal experiment/ (1275917)

42	animal experiment/ not (human experiment/ or human/) (2685603)

43	or/41-42 (2763141)

44	2 not 43 (253)

45	2 and 43 (20)

46	4 (209)

47	7 (210)

48	14 (209)

49	21 (198)

50	24 (210)

51	27 (256)

52	30 (208)

53	33 (208)

54	36 (208)

55	39 (207)

56	44 (253)

### **Categories of Removed Records which should have been Retained:**

#### **1. Uses human participants or data**

Embase via Ovid http://ovidsp.ovid.com/

Database Segment and Indexing: <1974 to 2024 December 09>

Date Searched: 10^th^ December 2024

1	("2032685122" or "2032771169" or "638682177" or "628695725" or "627605583" or "2024814998" or "2016842768" or "2023737818" or "628240273" or "2021043886" or "626456781" or "2029517709" or "2032613524" or "2032454907" or "2030326502" or "2020587236" or "644440354" or "2032776813" or "2028593107" or "2032488833" or "2032395002" or "643071756" or "2028214291" or "2018155161" or "2031596371" or "2032941225" or "2021629501" or "2026945966" or "2025489109" or "2030896935" or "644241685" or "2028194233" or "623187887" or "2032914131" or "2029655544" or "2030262495" or "2017611704" or "2030243993" or "2021043299" or "2030284637" or "2028974500" or "2018760412" or "27261765" or "2030030570" or "2020587042" or "2021629005" or "2027603359" or "2030334400" or "643760085" or "643760086" or "2016843570" or "2018154786" or "2020191383" or "626477948" or "2030897464" or "2016899791" or "2029877847" or "2017166249" or "2032987836" or "2017166852" or "2030038908" or "2032354278" or "2032955412" or "2022548382" or "2017166776" or "2024359373" or "2016370733" or "2032068230" or "2024359322" or "2017166594" or "2001155219" or "2032789358" or "2024218404" or "2019150167" or "2020191438" or "2022075558" or "2020587370" or "2027603516" or "2020587003" or "644265847" or "2019603154" or "2030783419" or "644436004" or "2028460176" or "2032068772" or "2029633608" or "2028494494" or "2001421801" or "2015874017" or "644485422" or "2031058328" or "2028194510" or "624263703" or "644556004" or "2030246424" or "2029775041" or "2031394940" or "2026946050" or "2030187219" or "2030783620" or "2024395294" or "2027772510" or "627463941" or "2016897994" or "2017166051" or "2018694304" or "2032838781" or "2015874207" or "2016843452" or "2029562426" or "2023737693" or "2027192276" or "2030335951" or "2032069053" or "2030219820" or "625863316" or "2002179641" or "2029874938" or "2030038251" or "629494810" or "2027603136" or "644563349" or "2026946169" or "2030219558" or "2028194498" or "2028194858" or "626452684" or "2026945776" or "2032886317" or "2029905647" or "2018155143" or "644563580" or "2031082477" or "2030783458" or "2016843021" or "2023737603" or "2020675525" or "2018154693" or "2030727482" or "2025503404" or "2022075707" or "625667182" or "2001203560" or "626437797" or "2025488494" or "2028194406" or "2031605050" or "644056962" or "644561863" or "624563910" or "2030314957" or "70253318" or "2029653041" or "2015873897" or "643676676" or "2021629015" or "630319434" or "2030348166" or "2029623143" or "2017166650" or "2001493084" or "2029585738" or "2027603423" or "2031971346" or "2029891050" or "2021629296" or "643260981" or "2016843667" or "2023146671" or "624431117" or "2029635162" or "2032150753" or "2018694324" or "2030783992" or "2030956632" or "2032932470" or "644261081" or "2017166815" or "2032681611" or "2032937888" or "2001067459" or "2029821280" or "2032942919" or "2019150453" or "2018694217" or "2023737672" or "2027993188" or "2030219275" or "636478876" or "642818617" or "2001445386" or "644441798" or "46106352" or "2029877721" or "641654494" or "2026214994" or "2025488931" or "2027156881" or "2031394826" or "2017166262" or "2032068217" or "626435471" or "643574264" or "2024974806" or "2024975157" or "2032867352" or "2020586973" or "624564732" or "2032069048" or "2016843684" or "2030250761" or "2032939245" or "2026945678" or "2023147078" or "644242441" or "625343769" or "2032982601" or "2024359021" or "2031394896" or "2030338559" or "2027848002" or "2029879726" or "2023737235" or "2017611133" or "2029518029" or "2015874265" or "2018155381" or "2022075591" or "2027602581" or "2032867622" or "644427631" or "2029353042" or "2029291482" or "2021629526" or "611554644" or "644576312" or "2017166713" or "2004836513" or "2029800773" or "2030654945" or "2026946096" or "2022242632" or "2032429194" or "2001401036" or "2021628969" or "2030784054" or "629325082" or "2030285074" or "2030181271" or "2003529175" or "2031632352" or "644314905" or "2002953028" or "623322732" or "2029930833" or "2015511617" or "643566330" or "2005043535" or "2028194891" or "624714843" or "2026215414" or "2024755348" or "2025488794" or "2024975108" or "2032859207" or "2015039721" or "2015039229" or "2015873913" or "2019602655" or "2030061894" or "2023146808" or "2010153094" or "2031823453" or "630349504" or "633306989" or "2029646454" or "2018693964" or "2007786479" or "2018154590" or "2029646460" or "2032394965" or "2030285023" or "2016842997" or "2029895436" or "2027950551" or "2032452450" or "2030163948" or "2029867873" or "623154909" or "2029646427" or "610669609" or "2031394660" or "2032859001" or "643906291" or "2032720124" or "2015874126" or "2018694471" or "350156070" or "644314834" or "2029455211" or "2032652581" or "2030173443" or "644314802" or "2032884476" or "2022075222" or "2025488747" or "2022074225" or "2029448005" or "2029518419" or "644588971" or "2032047497" or "624431615" or "2032953633" or "2001401788" or "2030314912" or "2017611020" or "624563880" or "2030274502" or "2018215835" or "2023147235" or "623173687" or "2029273157" or "2031006418" or "2030342098" or "2030783474" or "2018694548" or "2029565031" or "2030784294" or "638826889" or "2021629409" or "2030348675" or "2020587193" or "2030231115" or "2028491452" or "2032866774" or "2018154825" or "2032380475" or "2026892686" or "2004572601" or "642033997" or "2019150169" or "2030303340" or "2032821807" or "644555931" or "2030235843" or "2032393866" or "2024974649" or "2030199399" or "2017611846" or "2022548175" or "2026946475" or "2029877697" or "627237353" or "644257607" or "2024359064" or "2030342196" or "2016370415" or "644576385" or "625572742" or "2017611156" or "2031393997" or "626478658" or "2031394010" or "2023738813" or "2030243854" or "2031394509" or "623298294" or "2028779274" or "644586386" or "644556730" or "644440855" or "644586392" or "644439409" or "2031394019" or "627464487" or "2020191906" or "2032811133" or "2011263619" or "2023328117" or "2024975058" or "2015511535" or "2032346087" or "2031005101" or "625343923" or "2032855027" or "2027603402" or "2024359119" or "2030266713" or "2017677574" or "2031752330" or "2026215133" or "2032896805" or "2030322701" or "2029517713" or "2026945975" or "2027952223" or "644283613" or "2001445858" or "2029211348" or "2016842966" or "2019150523" or "2026290018" or "2026215408" or "2030322350" or "639393771" or "2014023305" or "2022075252" or "2030073163" or "2017611365" or "2022075667" or "629427814" or "2024975180" or "51889046" or "2026890439" or "2029140317" or "2030250452" or "2032996730" or "2015038998" or "2029869036" or "2029870186" or "644534549" or "2030270895" or "2030219575" or "2031386474" or "2031089068" or "627191017" or "2023737204" or "2032492463" or "2032613525" or "2028194225" or "2032418544" or "2030342063" or "2031634533" or "2030905072" or "2032069029" or "2029466981" or "624431246" or "2032560950" or "2030783490" or "2016843444" or "2032821989" or "644314070" or "2026215337" or "2019770967" or "2019603011" or "2016843405" or "2030303390" or "620982075" or "626434580" or "2029518043" or "2026945707" or "2017678969" or "2017611594" or "2023737483" or "644284071" or "2030784274" or "629337866" or "2020223710" or "626460770" or "2024975256" or "2022548080" or "2020191306" or "2029518274" or "2026946041" or "2026889279" or "2024174492" or "2032859214" or "623187625" or "2030219873" or "2028194508" or "50745671" or "2032068313" or "2002912198" or "2031446121" or "2028036998" or "2030831756" or "2030226531" or "2001808382" or "2032068138" or "2017611884" or "2020587768" or "2002078048" or "2029517994" or "2031135166" or "2022548540" or "632053853" or "2016370405" or "2022548313" or "2031327700" or "2032299368" or "644314565" or "2032939674" or "2030783912" or "2032838778" or "2032775239" or "2029730089" or "2029871403" or "2015873743" or "2030284565" or "2022693608" or "2004611240" or "2030340002" or "2030246907" or "2026490434" or "2017611189" or "608115609" or "2031817883" or "2022547822" or "2024974702" or "627251485" or "2030274550" or "2024397358" or "2032433852" or "644314162" or "2032047715" or "2001405459" or "2032774897" or "2029517998" or "2029603363" or "2022075089" or "2017611851" or "2013951939" or "636170043" or "2028194144" or "2030783704" or "2018154685" or "2020191579" or "2018694284" or "2029809136" or "2029258990" or "604426625" or "2032469465" or "2032986354" or "2021628919" or "628695947" or "2022870539" or "2028778615" or "2015873929" or "624431756" or "2016370885" or "29467080" or "2024974930" or "2029529086" or "2032623800" or "2029517964" or "644514480" or "2032914907" or "629410794" or "626516921" or "2028779222" or "2027187311" or "625977407" or "2029347975" or "2019150489" or "627190366" or "644522222" or "2017611595" or "2018154747" or "2029517783" or "2030244223" or "2034394337" or "2030248317" or "2026214975" or "2032392197" or "2022115046" or "2017610866" or "2029935100" or "2017166143" or "643997978" or "2031634594" or "2017166284" or "624431310" or "2019770681" or "2032601537" or "630320419" or "2032910911" or "2032382905" or "2029175727" or "2018155067" or "2030220067" or "2029517622" or "2028644292" or "2030303425" or "2028778797" or "2028779352" or "2017611212" or "2032369817" or "630050130" or "2032068721" or "2030219675" or "2031183114" or "2023147229" or "2029775006" or "70088192" or "2028195100" or "644344183" or "2027602611" or "629302343" or "615253305" or "644435842" or "629411398" or "2029874481" or "2025000686" or "628695406" or "2032819796" or "2018155232" or "2032861656" or "2027603001" or "624732960" or "2031394150" or "2023737319" or "2030303156" or "2029518335" or "636275965" or "2019150225" or "624564924" or "625347283" or "2019769273" or "2026945774" or "2030274563" or "644441232" or "644269645" or "2025488559" or "2029421830" or "2020191637" or "2029517887" or "2032875800" or "2021629038" or "2021629577" or "2017166413" or "2031175758" or "643567614" or "2026180049" or "2030397400" or "2030219868" or "7113283" or "2016030860" or "2016370511" or "2032891019" or "2016371240" or "644266176" or "624431280" or "2020192017" or "2028928211" or "2026319565" or "623154921" or "2030336357" or "644562503" or "2032211358" or "2026215051" or "2027785892" or "644313859" or "2029748237" or "2030326893" or "2023737562" or "2026945643" or "2023737335" or "2029517941" or "2028427482" or "2030262901" or "2022548526" or "640429090" or "2014062030" or "2032844530" or "2032111170" or "2032958009" or "643508604" or "2003436349" or "2017158748" or "642789249" or "2032273739" or "2021629433" or "2016370427" or "2030219687" or "2015874278" or "644242237" or "2026945543" or "2027602791" or "2019150395" or "2028195018" or "644314068" or "2019602712" or "2005627209" or "2022075622" or "2028194523" or "624431383" or "2032188827" or "644242186" or "624563897" or "2032884331" or "2020191590" or "2012993679" or "2031394652" or "2030783894" or "2013542514" or "643676170" or "643676409" or "643676777" or "2013601213" or "617374161" or "2032923850" or "2019150406" or "2029703044" or "2030285176" or "2029175726" or "2021043326" or "2027603030" or "2015873827" or "2016843749" or "2018693988" or "2030284645" or "2032800038" or "2028778606" or "2015896454" or "2030257601" or "2030326659" or "2015874336" or "631118405" or "2031002182" or "2031596815" or "2028194717" or "2019149777" or "2032879228" or "643364125" or "2025489014" or "2025489013" or "2019150107" or "2028014487" or "2031673801" or "2032953590" or "2032902124" or "2030207206" or "2029481362" or "643676774" or "2032068856").an. (735)

2	remove duplicates from 1 (735)

3	(animal/ or animal experiment/ or animal model/ or animal tissue/ or nonhuman/) not exp human/ (7145036)

4	2 not 3 (640)

5	2 and 3 (95)

6	(animal/ or nonhuman/) not exp human/ (6803417)

7	2 not 6 (640)

8	2 and 6 (95)

9	animal/ or exp animal experiment/ or nonhuman/ (9996171)

10	(rat or rats or mouse or mice or hamster or hamsters or animal or animals or dog or dogs or cat or cats or bovine or sheep).ti,sh. (5806446)

11	or/9-10 (10559138)

12	exp human/ or human experiment/ (27417348)

13	11 not (11 and 12) (7513136)

14	2 not 13 (640)

15	2 and 13 (95)

16	animal/ or exp animal experiment/ or nonhuman/ (9996171)

17	(rat or rats or mouse or mice or hamster or hamsters or animal or animals or dog or dogs or cat or cats or bovine or sheep).ti,ab,sh. (6799554)

18	or/16-17 (10969851)

19	exp human/ or human experiment/ (27417348)

20	18 not (18 and 19) (7650252)

21	2 not 20 (630)

22	2 and 20 (105)

23	((animal or nonhuman) not human).sh. (6805736)

24	2 not 23 (639)

25	2 and 23 (96)

26	animal experiment/ not (human experiment/ or human/) (2692876)

27	2 not 26 (655)

28	2 and 26 (80)

29	(exp animal/ or exp invertebrate/ or nonhuman/ or animal experiment/ or animal tissue/ or animal model/ or exp plant/ or exp fungus/) not (exp human/ or human tissue/) (8094699)

30	2 not 29 (638)

31	2 and 29 (97)

32	(exp animal/ or exp invertebrate/ or nonhuman/ or animal experiment/ or animal tissue/ or animal model/ or exp plant/ or exp fungus/) not (exp human/ or human tissue/ or human experiment/) (8093446)

33	2 not 32 (638)

34	2 and 32 (97)

35	(exp animal/ or exp invertebrate/ or nonhuman/ or animal experiment/ or animal tissue/ or animal model/) not (exp human/ or human tissue/ or human experiment/) (7595651)

36	2 not 35 (641)

37	2 and 35 (94)

38	(exp animals/ or exp animal experimentation/ or exp animal experiment/ or exp models animal/ or nonhuman/ or exp vertebrate/ or exp vertebrates/) not (exp humans/ or exp human experimentation/ or exp human experiment/) (7598106)

39	2 not 38 (639)

40	2 and 38 (96)

41	(rat or rats or mouse or mice or swine or porcine or murine or sheep or lambs or pigs or piglets or rabbit or rabbits or cat or cats or dog or dogs or cattle or bovine or monkey or monkeys or trout or marmoset$1).ti. and animal experiment/ (1278971)

42	animal experiment/ not (human experiment/ or human/) (2692876)

43	or/41-42 (2770986)

44	2 not 43 (633)

45	2 and 43 (102)

46	5 (95)

47	8 (95)

48	15 (95)

49	22 (105)

50	25 (96)

51	28 (80)

52	31 (97)

53	34 (97)

54	37 (94)

55	40 (96)

56	45 (102)

#### **2. Review or overview of a condition or intervention affecting humans**

Embase via Ovid http://ovidsp.ovid.com/

Database Segment and Indexing: <1974 to 2024 December 09>

Date Searched: 10^th^ December 2024

1	("636981787" or "613272064" or "2018019540" or "2029225400" or "2028886256" or "2013184318" or "2029937784" or "2030783574" or "2019936653" or "636312895" or "2030338787" or "2030672513" or "2017145036" or "2030834936" or "626052682" or "630485871" or "2013293812" or "2028503298" or "2022547860" or "2026811161" or "2031170465" or "2029214391" or "2024975241" or "2022075345" or "2032583027" or "2018694467" or "2032939635" or "2029058279" or "2004085360" or "632628017" or "628710879" or "618432880" or "2032839169" or "2026712696" or "2007638320" or "2018423270" or "2027734972" or "2007239241" or "2018839336" or "2025487593" or "2029863535" or "2012508457" or "2031393982" or "2008043484" or "2028059086" or "2031594003" or "2032111271" or "2029042375" or "644396776" or "2007035540" or "637040599" or "2023536854" or "2030783433" or "626912187" or "2029247586" or "2031889102" or "2022075841" or "644098429" or "629309720" or "2028965509" or "2032501626" or "2032867261" or "2030307050" or "2027348847" or "2028904923" or "2006923284" or "2005178890" or "2029646471" or "2021808000" or "2027948706" or "2030181076" or "2030727506" or "2016370739" or "2030074931" or "2027849811" or "2016958049" or "2017763483" or "2029688480" or "2013583403" or "29128277" or "2029161891" or "2029562135" or "2029422012" or "2027959924" or "2010048104" or "2032677946" or "2032146773" or "2025489243" or "2029441471" or "643564439" or "2007804685" or "2031394403" or "2029732206" or "2027113897" or "2031292174" or "71947033" or "2032894719" or "2028778829" or "2032669829" or "2029295086" or "2030033367" or "2018694449" or "2028261002" or "2030304063" or "2025856530" or "2015722460" or "2027798801" or "2032068906" or "643803020" or "2029066885" or "2023019688" or "2024359117" or "626758512" or "2026469789" or "624200522" or "2030153952" or "2029059180" or "2030221597" or "2029270720" or "2018839210" or "2030212203" or "635342521" or "644519809" or "2014112693" or "2014259992" or "629783491" or "2014157754" or "2032248300" or "2030932779" or "2006132160" or "2018154864" or "2032200741" or "600631152" or "6191273" or "7088453" or "2015874018" or "2024974636" or "2028841341" or "2032038663" or "2032915013" or "2032909485" or "627135574" or "2030143841" or "2021394476" or "2029894601" or "2029397074" or "2030104247" or "2029703009" or "2031182547" or "2030274988" or "2007173764" or "2027245256" or "613548076" or "2029413571" or "2013616422").an. (155)

2	remove duplicates from 1 (155)

3	(animal/ or animal experiment/ or animal model/ or animal tissue/ or nonhuman/) not exp human/ (7145036)

4	2 not 3 (97)

5	2 and 3 (58)

6	(animal/ or nonhuman/) not exp human/ (6803417)

7	2 not 6 (98)

8	2 and 6 (57)

9	animal/ or exp animal experiment/ or nonhuman/ (9996171)

10	(rat or rats or mouse or mice or hamster or hamsters or animal or animals or dog or dogs or cat or cats or bovine or sheep).ti,sh. (5806446)

11	or/9-10 (10559138)

12	exp human/ or human experiment/ (27417348)

13	11 not (11 and 12) (7513136)

14	2 not 13 (97)

15	2 and 13 (58)

16	animal/ or exp animal experiment/ or nonhuman/ (9996171)

17	(rat or rats or mouse or mice or hamster or hamsters or animal or animals or dog or dogs or cat or cats or bovine or sheep).ti,ab,sh. (6799554)

18	or/16-17 (10969851)

19	exp human/ or human experiment/ (27417348)

20	18 not (18 and 19) (7650252)

21	2 not 20 (86)

22	2 and 20 (69)

23	((animal or nonhuman) not human).sh. (6805736)

24	2 not 23 (98)

25	2 and 23 (57)

26	animal experiment/ not (human experiment/ or human/) (2692876)

27	2 not 26 (150)

28	2 and 26 (5)

29	(exp animal/ or exp invertebrate/ or nonhuman/ or animal experiment/ or animal tissue/ or animal model/ or exp plant/ or exp fungus/) not (exp human/ or human tissue/) (8094699)

30	2 not 29 (92)

31	2 and 29 (63)

32	(exp animal/ or exp invertebrate/ or nonhuman/ or animal experiment/ or animal tissue/ or animal model/ or exp plant/ or exp fungus/) not (exp human/ or human tissue/ or human experiment/) (8093446)

33	2 not 32 (92)

34	2 and 32 (63)

35	(exp animal/ or exp invertebrate/ or nonhuman/ or animal experiment/ or animal tissue/ or animal model/) not (exp human/ or human tissue/ or human experiment/) (7595651)

36	2 not 35 (96)

37	2 and 35 (59)

38	(exp animals/ or exp animal experimentation/ or exp animal experiment/ or exp models animal/ or nonhuman/ or exp vertebrate/ or exp vertebrates/) not (exp humans/ or exp human experimentation/ or exp human experiment/) (7598106)

39	2 not 38 (96)

40	2 and 38 (59)

41	(rat or rats or mouse or mice or swine or porcine or murine or sheep or lambs or pigs or piglets or rabbit or rabbits or cat or cats or dog or dogs or cattle or bovine or monkey or monkeys or trout or marmoset$1).ti. and animal experiment/ (1278971)

42	animal experiment/ not (human experiment/ or human/) (2692876)

43	or/41-42 (2770986)

44	2 not 43 (150)

45	2 and 43 (5)

46	5 (58)

47	8 (57)

48	15 (58)

49	22 (69)

50	25 (57)

51	28 (5)

52	31 (63)

53	34 (63)

54	37 (59)

55	40 (59)

56	45 (5)

#### **3. Review or overview of a factor affecting human health**

Embase via Ovid http://ovidsp.ovid.com/

Database Segment and Indexing: <1974 to 2024 December 09>

Date Searched: 10^th^ December 2024

1	("2029892367" or "2017166390" or "2031135945" or "2016816256" or "2030346026" or "2027972980" or "2029196909" or "2029965494" or "644564495" or "2030784229" or "2016370876" or "644510056" or "2014898143" or "2028194261" or "2032942967" or "2029149387" or "2028825860" or "2029635567" or "2017581891" or "2022075518" or "2031394512" or "2030826032" or "644586446" or "634932623" or "2031506511" or "2027944607" or "2028130533" or "2028997854" or "2028194526" or "2017611289" or "2027848981" or "2030007139" or "2030114419" or "2029771314" or "644587447" or "362270316" or "2029647007" or "2032150106" or "2025149147" or "2028194311" or "2016199852" or "2030783932" or "2011864415" or "2027734157" or "2001855108" or "2025173222" or "2018694238" or "2029742940" or "2032660776" or "2014389281" or "2017166619" or "2029741316" or "2032950631" or "2030783630" or "2021629300" or "2019602877").an. (56)

2	remove duplicates from 1 (56)

3	(animal/ or animal experiment/ or animal model/ or animal tissue/ or nonhuman/) not exp human/ (7145036)

4	2 not 3 (42)

5	2 and 3 (14)

6	(animal/ or nonhuman/) not exp human/ (6803417)

7	2 not 6 (43)

8	2 and 6 (13)

9	animal/ or exp animal experiment/ or nonhuman/ (9996171)

10	(rat or rats or mouse or mice or hamster or hamsters or animal or animals or dog or dogs or cat or cats or bovine or sheep).ti,sh. (5806446)

11	or/9-10 (10559138)

12	exp human/ or human experiment/ (27417348)

13	11 not (11 and 12) (7513136)

14	2 not 13 (42)

15	2 and 13 (14)

16	animal/ or exp animal experiment/ or nonhuman/ (9996171)

17	(rat or rats or mouse or mice or hamster or hamsters or animal or animals or dog or dogs or cat or cats or bovine or sheep).ti,ab,sh. (6799554)

18	or/16-17 (10969851)

19	exp human/ or human experiment/ (27417348)

20	18 not (18 and 19) (7650252)

21	2 not 20 (41)

22	2 and 20 (15)

23	((animal or nonhuman) not human).sh. (6805736)

24	2 not 23 (43)

25	2 and 23 (13)

26	animal experiment/ not (human experiment/ or human/) (2692876)

27	2 not 26 (54)

28	2 and 26 (2)

29	(exp animal/ or exp invertebrate/ or nonhuman/ or animal experiment/ or animal tissue/ or animal model/ or exp plant/ or exp fungus/) not (exp human/ or human tissue/) (8094699)

30	2 not 29 (40)

31	2 and 29 (16)

32	(exp animal/ or exp invertebrate/ or nonhuman/ or animal experiment/ or animal tissue/ or animal model/ or exp plant/ or exp fungus/) not (exp human/ or human tissue/ or human experiment/) (8093446)

33	2 not 32 (40)

34	2 and 32 (16)

35	(exp animal/ or exp invertebrate/ or nonhuman/ or animal experiment/ or animal tissue/ or animal model/) not (exp human/ or human tissue/ or human experiment/) (7595651)

36	2 not 35 (41)

37	2 and 35 (15)

38	(exp animals/ or exp animal experimentation/ or exp animal experiment/ or exp models animal/ or nonhuman/ or exp vertebrate/ or exp vertebrates/) not (exp humans/ or exp human experimentation/ or exp human experiment/) (7598106)

39	2 not 38 (41)

40	2 and 38 (15)

41	(rat or rats or mouse or mice or swine or porcine or murine or sheep or lambs or pigs or piglets or rabbit or rabbits or cat or cats or dog or dogs or cattle or bovine or monkey or monkeys or trout or marmoset$1).ti. and animal experiment/ (1278971)

42	animal experiment/ not (human experiment/ or human/) (2692876)

43	or/41-42 (2770986)

44	2 not 43 (54)

45	2 and 43 (2)

46	5 (14)

47	8 (13)

48	15 (14)

49	22 (15)

50	25 (13)

51	28 (2)

52	31 (16)

53	34 (16)

54	37 (15)

55	40 (15)

56	45 (2)

#### **4. Study outcomes relevant to diagnosing or treating humans**

Embase via Ovid http://ovidsp.ovid.com/

Database Segment and Indexing: <1974 to 2024 December 09>

Date Searched: 10^th^ December 2024

1	("2024291614" or "2007088885" or "2029840180" or "2028979836" or "2016080889" or "2001511851" or "2018694563" or "2020191876" or "2030173479" or "2019770506" or "2032720123" or "644243235" or "2029678228" or "2030784110" or "2020755740" or "2001672726" or "2020676082" or "631271087" or "2032452604" or "2030243934" or "644447459" or "2017613916" or "2007415928" or "2030784151" or "2029949483" or "620308849" or "2030025694" or "625666899" or "643676516").an. (29)

2	remove duplicates from 1 (29)

3	(animal/ or animal experiment/ or animal model/ or animal tissue/ or nonhuman/) not exp human/ (7145036)

4	2 not 3 (20)

5	2 and 3 (9)

6	(animal/ or nonhuman/) not exp human/ (6803417)

7	2 not 6 (20)

8	2 and 6 (9)

9	animal/ or exp animal experiment/ or nonhuman/ (9996171)

10	(rat or rats or mouse or mice or hamster or hamsters or animal or animals or dog or dogs or cat or cats or bovine or sheep).ti,sh. (5806446)

11	or/9-10 (10559138)

12	exp human/ or human experiment/ (27417348)

13	11 not (11 and 12) (7513136)

14	2 not 13 (20)

15	2 and 13 (9)

16	animal/ or exp animal experiment/ or nonhuman/ (9996171)

17	(rat or rats or mouse or mice or hamster or hamsters or animal or animals or dog or dogs or cat or cats or bovine or sheep).ti,ab,sh. (6799554)

18	or/16-17 (10969851)

19	exp human/ or human experiment/ (27417348)

20	18 not (18 and 19) (7650252)

21	2 not 20 (19)

22	2 and 20 (10)

23	((animal or nonhuman) not human).sh. (6805736)

24	2 not 23 (20)

25	2 and 23 (9)

26	animal experiment/ not (human experiment/ or human/) (2692876)

27	2 not 26 (28)

28	2 and 26 (1)

29	(exp animal/ or exp invertebrate/ or nonhuman/ or animal experiment/ or animal tissue/ or animal model/ or exp plant/ or exp fungus/) not (exp human/ or human tissue/) (8094699)

30	2 not 29 (18)

31	2 and 29 (11)

32	(exp animal/ or exp invertebrate/ or nonhuman/ or animal experiment/ or animal tissue/ or animal model/ or exp plant/ or exp fungus/) not (exp human/ or human tissue/ or human experiment/) (8093446)

33	2 not 32 (18)

34	2 and 32 (11)

35	(exp animal/ or exp invertebrate/ or nonhuman/ or animal experiment/ or animal tissue/ or animal model/) not (exp human/ or human tissue/ or human experiment/) (7595651)

36	2 not 35 (20)

37	2 and 35 (9)

38	(exp animals/ or exp animal experimentation/ or exp animal experiment/ or exp models animal/ or nonhuman/ or exp vertebrate/ or exp vertebrates/) not (exp humans/ or exp human experimentation/ or exp human experiment/) (7598106)

39	2 not 38 (20)

40	2 and 38 (9)

41	(rat or rats or mouse or mice or swine or porcine or murine or sheep or lambs or pigs or piglets or rabbit or rabbits or cat or cats or dog or dogs or cattle or bovine or monkey or monkeys or trout or marmoset$1).ti. and animal experiment/ (1278971)

42	animal experiment/ not (human experiment/ or human/) (2692876)

43	or/41-42 (2770986)

44	2 not 43 (27)

45	2 and 43 (2)

46	5 (9)

47	8 (9)

48	15 (9)

49	22 (10)

50	25 (9)

51	28 (1)

52	31 (11)

53	34 (11)

54	37 (9)

55	40 (9)

56	45 (2)

#### **5. Study outcomes relevant to a factor affecting human health**

Embase via Ovid http://ovidsp.ovid.com/

Database Segment and Indexing: <1974 to 2024 December 09>

Date Searched: 10^th^ December 2024

1	("2031148060" or "2027666977" or "624753299" or "2028838288" or "643070680" or "2026215084" or "2026191461" or "2028803323" or "2031334625" or "2030203039" or "2016438657" or "2029211376" or "2026320351" or "2028658183" or "2027750124" or "359262404" or "2023615273" or "2008490664" or "2029091360" or "2028075455" or "2026811171" or "2029621180" or "2026822821" or "2027778123" or "2029490307" or "2024974751" or "46902063" or "629405131" or "2028872376" or "623683500" or "2029221714" or "2029924182" or "9240656" or "2031291554" or "2032771772" or "2020674826" or "2029559690" or "2030571049" or "2018311827" or "2032435246" or "2028011077" or "2030521505" or "2028585449" or "2028037446" or "2031826923" or "2029489478" or "2017754218" or "637302637" or "2029646435" or "2031470894" or "2030020589" or "2032859227" or "2028423884" or "2029361295" or "2027603246" or "2029442073" or "2032492490" or "2032608001" or "625158468" or "2030284835" or "2029407880" or "2004291033" or "2031711326" or "2030235106" or "602587040" or "2010547152" or "2030236196" or "2032283215" or "2030174267" or "2029064315" or "643899444" or "2025002246" or "2016843031" or "2024748212" or "2015873751" or "644256000" or "2030219674" or "2029813254" or "2025689373" or "2019455869" or "2032213028" or "2029815075" or "2031594983" or "2023606587" or "2015548200" or "2025675471" or "2030895419" or "2032866540").an. (88)

2	remove duplicates from 1 (88)

3	(animal/ or animal experiment/ or animal model/ or animal tissue/ or nonhuman/) not exp human/ (7145036)

4	2 not 3 (40)

5	2 and 3 (48)

6	(animal/ or nonhuman/) not exp human/ (6803417)

7	2 not 6 (40)

8	2 and 6 (48)

9	animal/ or exp animal experiment/ or nonhuman/ (9996171)

10	(rat or rats or mouse or mice or hamster or hamsters or animal or animals or dog or dogs or cat or cats or bovine or sheep).ti,sh. (5806446)

11	or/9-10 (10559138)

12	exp human/ or human experiment/ (27417348)

13	11 not (11 and 12) (7513136)

14	2 not 13 (39)

15	2 and 13 (49)

16	animal/ or exp animal experiment/ or nonhuman/ (9996171)

17	(rat or rats or mouse or mice or hamster or hamsters or animal or animals or dog or dogs or cat or cats or bovine or sheep).ti,ab,sh. (6799554)

18	or/16-17 (10969851)

19	exp human/ or human experiment/ (27417348)

20	18 not (18 and 19) (7650252)

21	2 not 20 (29)

22	2 and 20 (59)

23	((animal or nonhuman) not human).sh. (6805736)

24	2 not 23 (40)

25	2 and 23 (48)

26	animal experiment/ not (human experiment/ or human/) (2692876)

27	2 not 26 (74)

28	2 and 26 (14)

29	(exp animal/ or exp invertebrate/ or nonhuman/ or animal experiment/ or animal tissue/ or animal model/ or exp plant/ or exp fungus/) not (exp human/ or human tissue/) (8094699)

30	2 not 29 (38)

31	2 and 29 (50)

32	(exp animal/ or exp invertebrate/ or nonhuman/ or animal experiment/ or animal tissue/ or animal model/ or exp plant/ or exp fungus/) not (exp human/ or human tissue/ or human experiment/) (8093446)

33	2 not 32 (38)

34	2 and 32 (50)

35	(exp animal/ or exp invertebrate/ or nonhuman/ or animal experiment/ or animal tissue/ or animal model/) not (exp human/ or human tissue/ or human experiment/) (7595651)

36	2 not 35 (38)

37	2 and 35 (50)

38	(exp animals/ or exp animal experimentation/ or exp animal experiment/ or exp models animal/ or nonhuman/ or exp vertebrate/ or exp vertebrates/) not (exp humans/ or exp human experimentation/ or exp human experiment/) (7598106)

39	2 not 38 (38)

40	2 and 38 (50)

41	(rat or rats or mouse or mice or swine or porcine or murine or sheep or lambs or pigs or piglets or rabbit or rabbits or cat or cats or dog or dogs or cattle or bovine or monkey or monkeys or trout or marmoset$1).ti. and animal experiment/ (1278971)

42	animal experiment/ not (human experiment/ or human/) (2692876)

43	or/41-42 (2770986)

44	2 not 43 (72)

45	2 and 43 (16)

46	5 (48)

47	8 (48)

48	15 (49)

49	22 (59)

50	25 (48)

51	28 (14)

52	31 (50)

53	34 (50)

54	37 (50)

55	40 (50)

56	45 (16)

#### **6. Not on animals/nonhumans or human health directly**

Embase via Ovid http://ovidsp.ovid.com/

Database Segment and Indexing: <1974 to 2024 December 09>

Date Searched: 10^th^ December 2024

1	("2026215134" or "2017679583" or "2019150445" or "2030048131" or "636632816" or "2032347707" or "2030047923" or "2030322589" or "2030322494" or "373252075" or "2029866880" or "2032735621" or "2030243735" or "2030064565" or "2032718273" or "634930622" or "643616638" or "354655095" or "2030221449" or "2010599488" or "2010599491" or "629412034").an. (22)

2	remove duplicates from 1 (22)

3	(animal/ or animal experiment/ or animal model/ or animal tissue/ or nonhuman/) not exp human/ (7145036)

4	2 not 3 (19)

5	2 and 3 (3)

6	(animal/ or nonhuman/) not exp human/ (6803417)

7	2 not 6 (19)

8	2 and 6 (3)

9	animal/ or exp animal experiment/ or nonhuman/ (9996171)

10	(rat or rats or mouse or mice or hamster or hamsters or animal or animals or dog or dogs or cat or cats or bovine or sheep).ti,sh. (5806446)

11	or/9-10 (10559138)

12	exp human/ or human experiment/ (27417348)

13	11 not (11 and 12) (7513136)

14	2 not 13 (19)

15	2 and 13 (3)

16	animal/ or exp animal experiment/ or nonhuman/ (9996171)

17	(rat or rats or mouse or mice or hamster or hamsters or animal or animals or dog or dogs or cat or cats or bovine or sheep).ti,ab,sh. (6799554)

18	or/16-17 (10969851)

19	exp human/ or human experiment/ (27417348)

20	18 not (18 and 19) (7650252)

21	2 not 20 (14)

22	2 and 20 (8)

23	((animal or nonhuman) not human).sh. (6805736)

24	2 not 23 (19)

25	2 and 23 (3)

26	animal experiment/ not (human experiment/ or human/) (2692876)

27	2 not 26 (21)

28	2 and 26 (1)

29	(exp animal/ or exp invertebrate/ or nonhuman/ or animal experiment/ or animal tissue/ or animal model/ or exp plant/ or exp fungus/) not (exp human/ or human tissue/) (8094699)

30	2 not 29 (19)

31	2 and 29 (3)

32	(exp animal/ or exp invertebrate/ or nonhuman/ or animal experiment/ or animal tissue/ or animal model/ or exp plant/ or exp fungus/) not (exp human/ or human tissue/ or human experiment/) (8093446)

33	2 not 32 (19)

34	2 and 32 (3)

35	(exp animal/ or exp invertebrate/ or nonhuman/ or animal experiment/ or animal tissue/ or animal model/) not (exp human/ or human tissue/ or human experiment/) (7595651)

36	2 not 35 (19)

37	2 and 35 (3)

38	(exp animals/ or exp animal experimentation/ or exp animal experiment/ or exp models animal/ or nonhuman/ or exp vertebrate/ or exp vertebrates/) not (exp humans/ or exp human experimentation/ or exp human experiment/) (7598106)

39	2 not 38 (19)

40	2 and 38 (3)

41	(rat or rats or mouse or mice or swine or porcine or murine or sheep or lambs or pigs or piglets or rabbit or rabbits or cat or cats or dog or dogs or cattle or bovine or monkey or monkeys or trout or marmoset$1).ti. and animal experiment/ (1278971)

42	animal experiment/ not (human experiment/ or human/) (2692876)

43	or/41-42 (2770986)

44	2 not 43 (21)

45	2 and 43 (1)

46	5 (3)

47	8 (3)

48	15 (3)

49	22 (8)

50	25 (3)

51	28 (1)

52	31 (3)

53	34 (3)

54	37 (3)

55	40 (3)

56	45 (1)

#### **7. Other**

Embase via Ovid http://ovidsp.ovid.com/

Database Segment and Indexing: <1974 to 2024 December 09>

Date Searched: 10^th^ December 2024

1	("2030057439" or "2032706468" or "2029857886").an. (3)

2	remove duplicates from 1 (3)

3	(animal/ or animal experiment/ or animal model/ or animal tissue/ or nonhuman/) not exp human/ (7145036)

4	2 not 3 (3)

5	2 and 3 (0)

6	(animal/ or nonhuman/) not exp human/ (6803417)

7	2 not 6 (3)

8	2 and 6 (0)

9	animal/ or exp animal experiment/ or nonhuman/ (9996171)

10	(rat or rats or mouse or mice or hamster or hamsters or animal or animals or dog or dogs or cat or cats or bovine or sheep).ti,sh. (5806446)

11	or/9-10 (10559138)

12	exp human/ or human experiment/ (27417348)

13	11 not (11 and 12) (7513136)

14	2 not 13 (3)

15	2 and 13 (0)

16	animal/ or exp animal experiment/ or nonhuman/ (9996171)

17	(rat or rats or mouse or mice or hamster or hamsters or animal or animals or dog or dogs or cat or cats or bovine or sheep).ti,ab,sh. (6799554)

18	or/16-17 (10969851)

19	exp human/ or human experiment/ (27417348)

20	18 not (18 and 19) (7650252)

21	2 not 20 (3)

22	2 and 20 (0)

23	((animal or nonhuman) not human).sh. (6805736)

24	2 not 23 (3)

25	2 and 23 (0)

26	animal experiment/ not (human experiment/ or human/) (2692876)

27	2 not 26 (3)

28	2 and 26 (0)

29	(exp animal/ or exp invertebrate/ or nonhuman/ or animal experiment/ or animal tissue/ or animal model/ or exp plant/ or exp fungus/) not (exp human/ or human tissue/) (8094699)

30	2 not 29 (3)

31	2 and 29 (0)

32	(exp animal/ or exp invertebrate/ or nonhuman/ or animal experiment/ or animal tissue/ or animal model/ or exp plant/ or exp fungus/) not (exp human/ or human tissue/ or human experiment/) (8093446)

33	2 not 32 (3)

34	2 and 32 (0)

35	(exp animal/ or exp invertebrate/ or nonhuman/ or animal experiment/ or animal tissue/ or animal model/) not (exp human/ or human tissue/ or human experiment/) (7595651)

36	2 not 35 (3)

37	2 and 35 (0)

38	(exp animals/ or exp animal experimentation/ or exp animal experiment/ or exp models animal/ or nonhuman/ or exp vertebrate/ or exp vertebrates/) not (exp humans/ or exp human experimentation/ or exp human experiment/) (7598106)

39	2 not 38 (3)

40	2 and 38 (0)

41	(rat or rats or mouse or mice or swine or porcine or murine or sheep or lambs or pigs or piglets or rabbit or rabbits or cat or cats or dog or dogs or cattle or bovine or monkey or monkeys or trout or marmoset$1).ti. and animal experiment/ (1278971)

42	animal experiment/ not (human experiment/ or human/) (2692876)

43	or/41-42 (2770986)

44	2 not 43 (3)

45	2 and 43 (0)

46	5 (0)

47	8 (0)

48	15 (0)

49	22 (0)

50	25 (0)

51	28 (0)

52	31 (0)

53	34 (0)

54	37 (0)

55	40 (0)

56	45 (0)
